# Thrombosis and thrombocytopenia after vaccination against and infection with SARS-CoV-2: a population-based cohort analysis

**DOI:** 10.1101/2021.07.29.21261348

**Authors:** Edward Burn, Xintong Li, Antonella Delmestri, Nathan Jones, Talita Duarte-Salles, Carlen Reyes, Eugenia Martinez-Hernandez, Edelmira Marti, Katia MC Verhamme, Peter R Rijnbeek, Victoria Y Strauss, Daniel Prieto-Alhambra

**Author notes:** Corresponding author: Prof Daniel Prieto-Alhambra, Botnar Research Centre, Windmill Road, OX37LD, Oxford, UK. Joint first authors (Edward Burn, Xintong Li). Joint senior authors (Victoria Y Strauss, Daniel Prieto-Alhambra).

## Abstract

**Objectives:** To calculate the observed rates of thrombosis and thrombocytopenia following vaccination against SARS-CoV-2, infection with SARS-CoV-2, and to compare them to background (expected) rates in the general population.

**Design:** Cohort study using routinely collected primary care records.

**Setting:** Routine practice in the United Kingdom.

**Participants:** Two mutually exclusive vaccinated cohorts included people vaccinated with either ChAdOx1 or BNT162b2 between 8 December 2020 and 6 March 2021. A third cohort consisted of people newly infected with SARS-Cov-2 identified by a first positive RT-PCR test between 1 September 2020 and 28 February 2021. The fourth general population cohort for background rates included those people with a visit between 1 January 2017 and 31 December 2019. In total, we included 1,868,767 ChAdOx1 and 1,661,139 BNT162b2 vaccinees, 299,311 people infected with SARS-CoV-2, and 2,290,537 people from the general population.

**Interventions:** First-dose of either ChAdOx1 or BNT162b2

**Main outcome measures:** Outcomes included venous thrombosis, arterial thrombosis, thrombocytopenia, and thrombosis with thrombocytopenia. Outcome rates were estimated for recipients of the ChAdOx1 or BNT162b2 vaccines, for people infected with SARS-CoV-2, and background rates in the general population. Indirectly standardized incidence ratios (SIR) were estimated.

**Results:** We included 1,868,767 ChAdOx1 and 1,661,139 BNT162b2 vaccinees, 299,311 people infected with SARS-CoV-2, and 2,290,537 people from the general population for background rates. The SIRs for pulmonary embolism were 1.23 [95% CI, 1.09-1.39] after vaccination with ChAdOx1, 1.21 [1.07-1.36] after vaccination with BNT162b2, and 15.31 [14.08 to 16.65] for infection with SARS-CoV-2. The SIRs for thrombocytopenia after vaccination were 1.25 [1.19 to 1.31] for ChAdOx1 and 0.99 (0.94 to 1.04) for BNT162b2. Rates of deep vein thrombosis and arterial thrombosis were similar among those vaccinated and the general population.

**Conclusions:** ChAdOx1 and BNT162b2 had broadly similar safety profiles. Thrombosis rates after either vaccine were mostly similar to those of the general population. Rates of pulmonary embolism increased 1.2-fold after either vaccine and 15-fold with SARS-CoV-2 infection. Thrombocytopenia was more common among recipients of ChAdOx1 but not of BNT162b2.

**Summary box:** *What is already known on this topic:*

- Spontaneous reports of unusual and severe thrombosis with thrombocytopenia syndrome (TTS) raised concerns regarding the safety of adenovirus-based vaccines against SARS-CoV-2
- In a cohort study including over 280,000 people aged 18-65 years vaccinated with ChAdOx1 in Denmark and Norway, Pottegård et al reported increased rates of venous thromboembolic events as well as thrombocytopenia among vaccine recipients.

*What this study adds:*

- In this cohort study, ChAdOx1 and BNT162b2 were seen to have broadly similar safety profiles.
- Rates of thrombosis after either vaccine were generally similar to those of the general population. Rates of pulmonary embolism were though 1.2-fold higher than background rates after either vaccine, which compared to 15-fold higher after SARS-CoV-2 infection.
- Thrombocytopenia was more common among recipients of ChAdOx1 but not of BNT162b2.

## Introduction

Vaccines against SARS-CoV-2 have been developed rapidly using a number of platforms. The ChAdOx1 nCoV-19 (Oxford–AstraZeneca; ChAdOx1) and BNT162b2 mRNA (Pfizer– BioNTech; BNT162b2) vaccines received approval for use in the United Kingdom on 8 and 31 December 2020, respectively. Evidence from clinical trials and real-world data has shown these vaccines to be highly effective in preventing symptomatic COVID-19, severe disease, and hospitalization.^1–7^

As COVID-19 vaccines have been approved under emergency authorization, they must continue to be monitored to assess their safety. Instances of rare adverse events have been identified alongside the ongoing nationwide immunization programs.^8–10^ A particular concern has arisen regarding thrombotic events, with concurrent thrombocytopenia reported among individuals vaccinated with adenovirus-based vaccines against SARS-CoV-2. As of 26 May 2021, 348 spontaneous reports of major thromboembolic events with thrombocytopenia had been documented following 24 million first doses and 13 million second doses of the ChAdOx1 vaccine in the UK.^11^ Although fewer concerns have been raised about safety signals for BNT162b2, instances of immune thrombocytopenia have also been observed among recipients of this vaccine.^12^

In this study, we estimated the incidence of thrombosis, thrombocytopenia, and thrombosis with thrombocytopenia over the 28 days following a first dose of the ChAdOx1 and BNT162b2 vaccines and compared these rates with historical, pre-pandemic rates in the general population. To provide additional context, we also studied the rates of these events after a positive RT-PCR test for SARS-CoV-2.

## Methods

### Study design, setting, and data sources

People vaccinated against SARS-CoV-2 and people infected with SARS-CoV-2 were identified from Clinical Practice Research Datalink (CPRD) AURUM. A background cohort was obtained from CPRD GOLD to estimate pre-pandemic background rates. AURUM and GOLD are established primary care databases broadly representative of the UK population,^13, 14^ and previous research has demonstrated their validity for vaccine safety surveillance.^15, 16^ Both databases were mapped to the Observational Medical Outcomes Partnership (OMOP) Common Data Model (CDM) for analysis.^17^

### Study participants and follow-up

Four cohorts were studied. Two mutually exclusive vaccinated cohorts included people vaccinated with either ChAdOx1 or BNT162b2 between 8 December 2020 and 6 March 2021. They were followed for up to 28 days from their first vaccination (index date). A third cohort consisted of people newly infected with SARS-Cov-2 identified by a first positive RT-PCR test between 1 September 2020 and 28 February 2021. The test date was used as the index date. They were followed for up to 90 days. The fourth cohort, a general population background cohort, included people who had a primary care visit or contact recorded between 1 January 2017 and 31 December 2019. That first visit or contact was used as the index date and follow-up ran up to 31 December 2019.

All participants were required to be aged 30 years or older and, for the primary analysis, have at least 1 year of prior history available. Participants did not contribute to an analysis if they had the same event recorded in the year before their index date. Time at risk was censored if an individual had the outcome of interest or exited the database before the end of follow-up.

Sensitivity analyses were conducted removing the requirements of a year of prior history and, for the background population, a primary care visit or contact. For the latter, 1 January 2017 was used as the index date.

### Study outcomes

We used diagnostic codes to identify five venous thromboembolic events: cerebral venous sinus thrombosis (CVST), deep vein thrombosis (DVT), pulmonary embolism (PE), splanchnic vein thrombosis (SVT), and the composite event venous thromboembolism (VTE), which encompassed DVT and PE. We identified two arterial thromboembolic events (ATE): myocardial infarction and ischemic stroke. We also used an overall stroke definition that included non-specific, hemorrhagic, and ischemic stroke codes.

Thrombocytopenia was identified using diagnostic codes and laboratory data showing platelets count between 10,000 and 150,000 platelets per microliter, based on the Brighton collaboration definition.^18^ Immune thrombocytopenia was identified using diagnostic codes.

Thrombosis with thrombocytopenia syndromes (TTS) was identified where thrombocytopenia was observed within 10 days before or after thrombosis. This time window was broadened in a sensitivity analysis.

Results for additional related outcomes (intestinal infarction, platelet disorder, portal vein thrombosis, and thrombocytopenic purpura) are reported in the Appendix.

### Statistical methods

For each cohort and outcome of interest, we describe the cohort’s age, sex, comorbidities, and medication use within 6 months before and up to 4 days before the index date. We report the number of events observed and crude incidence rates per 100,000 person-years with 95% confidence intervals (CIs). We used indirect standardization with the background cohort as the standard to estimate the number of events expected for the vaccination and SARS-CoV-2 cohorts if their risk was the same as that of the general population.^19^ We estimated standardized incidence ratios (SIRs) and 95% confidence intervals comparing observed and expected rates. We stratified all analyses by 10-year age bands and sex and analyses of those vaccinated by calendar month. We calculated the standardized event difference proportion to provide a measure of absolute risk. To avoid re-identification, we do not report any analysis with under 5 cases.

The study protocol was published in the EU PAS Register (EUPAS40414). The use of CPRD data was approved by the Independent Scientific Advisory Committee (21_000391 and 20_000211). All analytical code is available at: https://github.com/oxford-pharmacoepi/CovidVaccinationSafetyStudy.

### Patient and public involvement

No patients or members of the public were directly involved in the study design or its execution.

## Results

We included 1,868,767 people vaccinated with ChAdOx1, 1,661,139 people vaccinated with BNT162b2, 299,311 people infected with SARS-CoV-2, and 2,290,537 people from the general population. The two vaccinated populations were similar in terms of age, sex, and clinical characteristics The vaccinated populations were older, more often female, and had a higher prevalence of all studied comorbidities than the general population. Differences in the prevalence of comorbidities was more pronounced for younger age groups. Those infected with SARS-CoV-2 were younger than the general population. Detailed characteristics for all four cohorts are given in Table 1, and stratified by age in the Appendix. BNT162b2 vaccination started earlier than ChAdOx1 vaccination. The numbers vaccinated with the two vaccines became similar in late January 2021, and ChAdOx1 vaccines predominated in February and early March (Figure 1).

**Figure 1.**
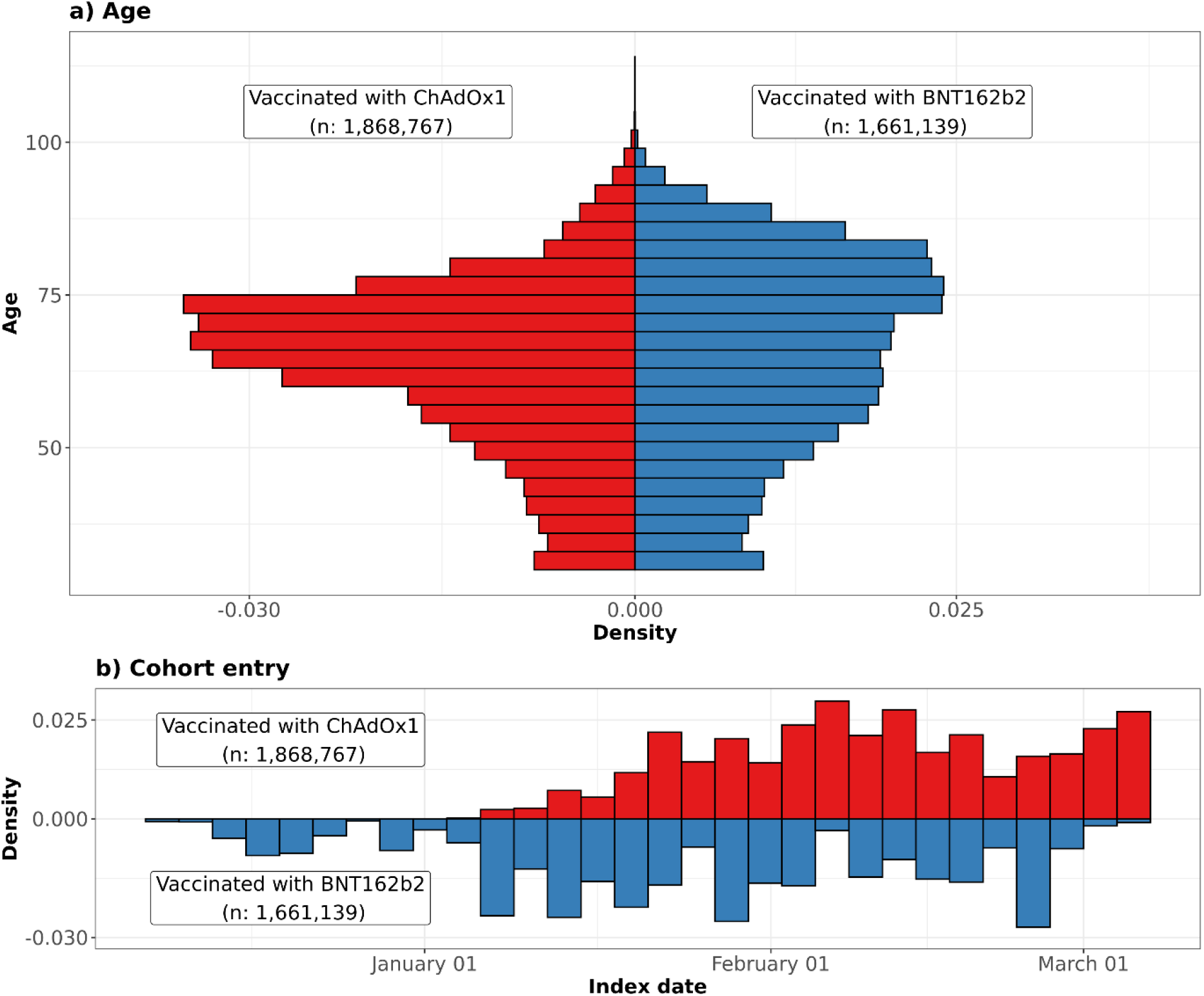
**Distribution of age profiles and date of cohort entry among people vaccinated against SARS-CoV-2**

**Table 1.** Characteristics of study participants. Characteristics of the participants in the four study cohorts used for the primary analyses. Participants were aged 30 years or older and had at least one year of prior history before index date in the database. Those in the general population had a primary care visit or contact between 2017 and 2019. People infected with SARS-CoV-2 had a confirmatory positive RT-PCR test. *Conditions of interest: autoimmune disease, antiphospholipid syndrome, thrombophilia, asthma, atrial fibrillation, malignant neoplastic disease, diabetes mellitus, obesity, or renal impairment. ^†^ Medications of interest: non-steroidal anti-inflammatory drugs, Cox2 inhibitors, systemic corticosteroids, hormonal contraceptives, tamoxifen, and sex hormones and modulators of the genital system.

|  | General population | Vaccinated with ChadOx1 | Vaccinated with BNT162b2 | Infected with SARS-CoV-2 |
| --- | --- | --- | --- | --- |
| N | 2,290,537 | 1,868,767 | 1,661,139 | 299,311 |
| Age (median [IQR]) | 54 [43 to 67] | 66 [56 to 73] | 67 [54 to 78] | 48 [39 to 58] |
| Age: 30 to 39 | 431,801 (18.9%) | 123,991 (6.6%) | 135,101 (8.1%) | 82,427 (27.5%) |
| Age: 40 to 49 | 459,400 (20.1%) | 173,453 (9.3%) | 178,503 (10.7%) | 77,184 (25.8%) |
| Age: 50 to 59 | 506,515 (22.1%) | 285,938 (15.3%) | 279,223 (16.8%) | 75,543 (25.2%) |
| Age: 60 to 69 | 403,673 (17.6%) | 567,041 (30.3%) | 322,252 (19.4%) | 38,646 (12.9%) |
| Age: 70 to 79 | 301,179 (13.1%) | 545,251 (29.2%) | 372,426 (22.4%) | 14,839 (5.0%) |
| Age: 80 or older | 187,969 (8.2%) | 173,093 (9.3%) | 373,634 (22.5%) | 10,672 (3.6%) |
| Sex: Male | 1,094,911 (47.8%) | 845,908 (45.3%) | 702,863 (42.3%) | 136,301 (45.5%) |
| Years of prior observation time (median [IQR]) | 13.5 [8.1 to 16.3] | 19.0 [7.7 to 31.1] | 19.1 [7.8 to 31.2] | 12.4 [5.3 to 23.9] |
| **Comorbidities** |  |  |  |  |
| Autoimmune disease | 57,773 (2.5%) | 72,119 (3.9%) | 64,311 (3.9%) | 7,136 (2.4%) |
| Antiphospholipid syndrome | 1,046 (0.0%) | 1,535 (0.1%) | 1,349 (0.1%) | 215 (0.1%) |
| Thrombophilia | 2,582 (0.1%) | 3,628 (0.2%) | 3,192 (0.2%) | 549 (0.2%) |
| Asthma | 301,669 (13.2%) | 284,126 (15.2%) | 253,177 (15.2%) | 45,470 (15.2%) |
| Atrial fibrillation | 72,643 (3.2%) | 100,730 (5.4%) | 117,398 (7.1%) | 5,993 (2.0%) |
| Malignant neoplastic disease | 187,499 (8.2%) | 257,449 (13.8%) | 268,655 (16.2%) | 15,663 (5.2%) |
| Diabetes mellitus | 197,619 (8.6%) | 306,944 (16.4%) | 276,194 (16.6%) | 28,488 (9.5%) |
| Obesity | 94,054 (4.1%) | 120,052 (6.4%) | 98,900 (6.0%) | 16,791 (5.6%) |
| Heart disease | 251,708 (11.0%) | 334,469 (17.9%) | 349,669 (21.0%) | 24,511 (8.2%) |
| Hypertensive disorder | 533,750 (23.3%) | 677,743 (36.3%) | 643,345 (38.7%) | 53,921 (18.0%) |
| Renal impairment | 161,596 (7.1%) | 207,751 (11.1%) | 234,386 (14.1%) | 14,150 (4.7%) |
| Chronic Obstructive<br>Pulmonary Disease | 77,205 (3.4%) | 107,636 (5.8%) | 95,654 (5.8%) | 5,384 (1.8%) |
| Dementia | 29,376 (1.3%) | 38,589 (2.1%) | 34,829 (2.1%) | 4,886 (1.6%) |
| ***Medication use (183 days prior to four days prior)*** |  |  |  |  |
| Non-steroidal anti-inflammatory drugs | 618,552 (27.0%) | 264,090 (14.1%) | 242,309 (14.6%) | 33,299 (11.1%) |
| Cox2 inhibitors | 6,115 (0.3%) | 1,931 (0.1%) | 1,581 (0.1%) | 258 (0.1%) |
| Systemic corticosteroids | 263,488 (11.5%) | 119,766 (6.4%) | 105,977 (6.4%) | 14,168 (4.7%) |
| Antithrombotic and anticoagulant therapies | 102,605 (4.5%) | 61,322 (3.3%) | 65,237 (3.9%) | 3,885 (1.3%) |
| Lipid modifying agents | 130,264 (5.7%) | 110,329 (5.9%) | 102,835 (6.2%) | 8,224 (2.7%) |
| Antineoplastic and immunomodulating agents | 57,403 (2.5%) | 20,445 (1.1%) | 20,541 (1.2%) | 3,822 (1.3%) |
| Hormonal contraceptives for systemic use | 76,796 (3.4%) | 18,566 (1.0%) | 20,491 (1.2%) | 7,205 (2.4%) |
| Tamoxifen | 1,957 (0.1%) | 814 (0.0%) | 799 (0.0%) | 87 (0.0%) |
| Sex hormones and modulators of the genital system | 110,583 (4.8%) | 44,612 (2.4%) | 44,563 (2.7%) | 10,283 (3.4%) |
| **Summary count of conditions and medications of interest** |  |  |  |  |
| One or more condition of interest* | 579,628 (25.3%) | 778,559 (41.7%) | 751,224 (45.2%) | 68,879 (23.0%) |
| One or more medication of interest<sup>†</sup> | 758,348 (33.1%) | 345,875 (18.5%) | 318,704 (19.2%) | 47,239 (15.8%) |
| One or more condition/medication of interest*<sup>†</sup> | 1,058,141 (46.2%) | 949,193 (50.8%) | 897,757 (54.0%) | 99,727 (33.3%) |

Rates of VTE were higher in men aged 50-59 vaccinated with BNT162b2 (367.5/100,000 person-years [95%CI 237.8-542.5]) than the age-sex specific background rate (211.9/100,000 person-years [200.2-224.1]) (Figure 2). Rates of VTE were also higher in women aged ≥80 vaccinated with ChAdOx1 (878.1/100,000 person-years [685.8-1107.6]) than the equivalent age-sex background (703.0/100,000 person-years [670.1-737.2]).

**Figure 2.**
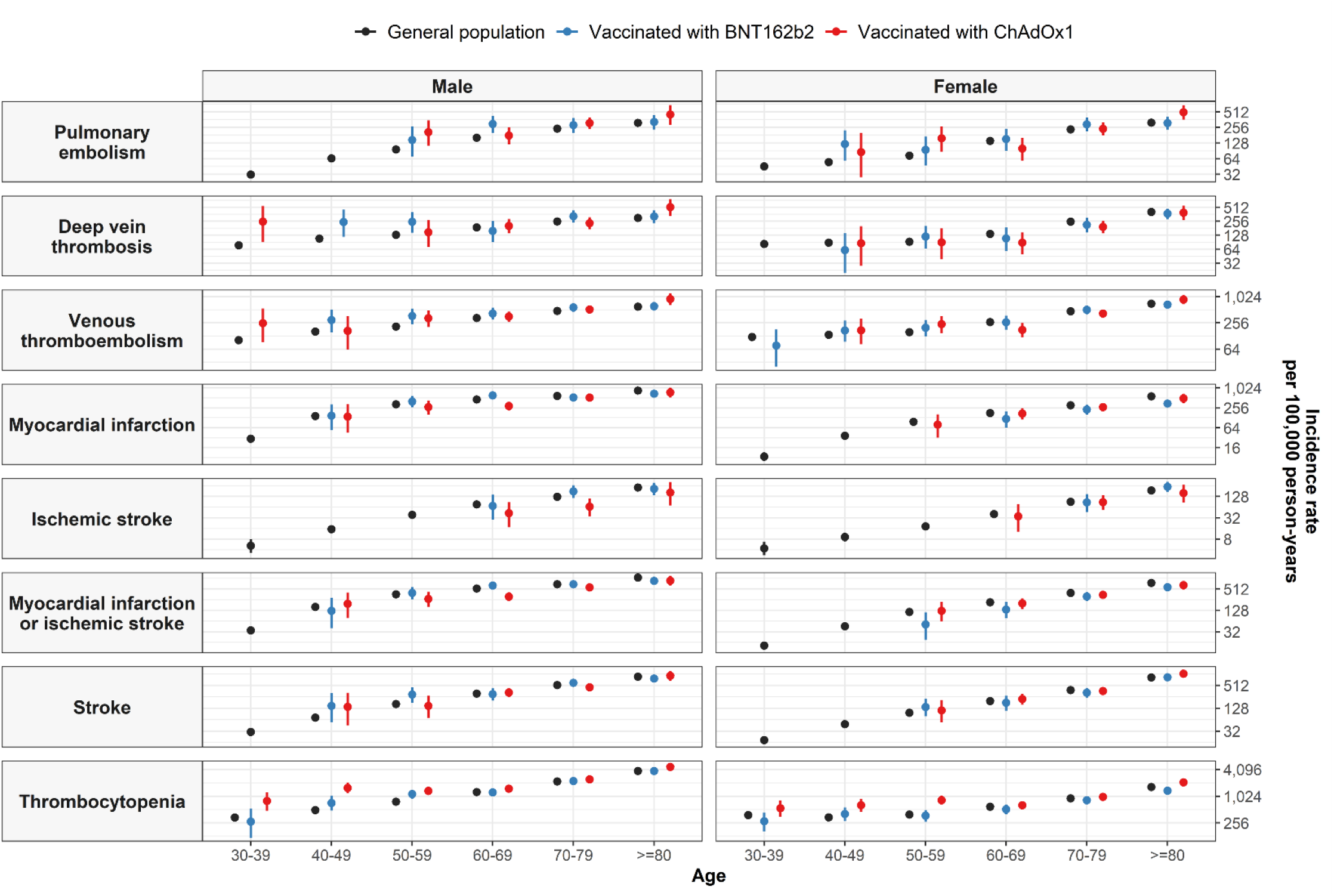
**Background and post-vaccine rates of thromboembolic events and thrombocytopenia by age and sex Events with less than 5 occurrences have been omitted for privacy reasons.**

Standardized SIRs for VTE were 1.07 [0.98-1.18] for ChAdOx1, 1.09 [1.00-1.20] for BNT162b2, and 8.08 [7.48-8.72] for SARS-Cov-2 (Table 2, Figure 3).

**Figure 3.**
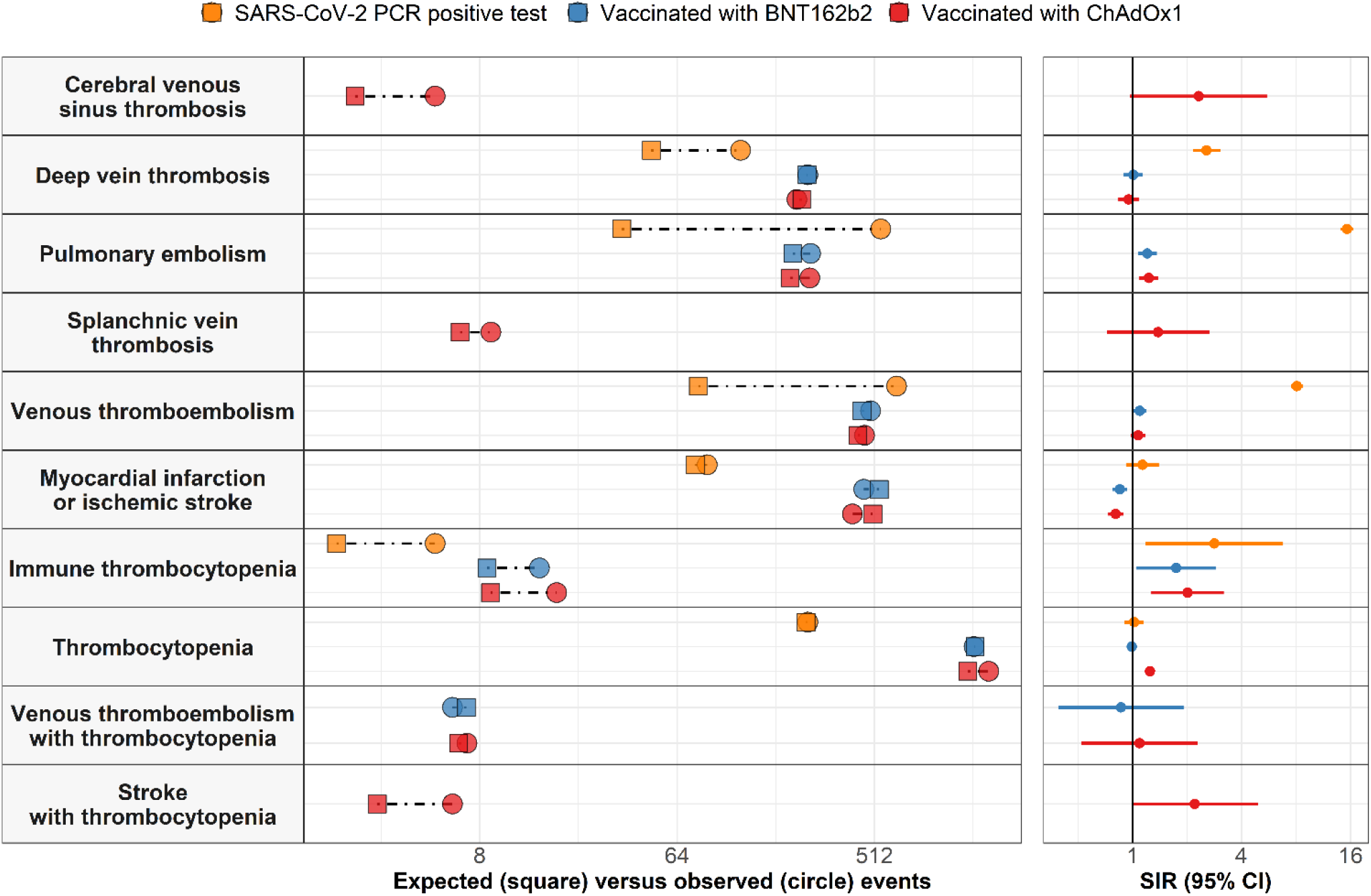
Expected versus observed events among those vaccinated against SARS-CoV-2 and those with a SARS-CoV-2 infection. Expected events for each of the study cohorts based on indirect standardization using rates from the general population between 2017 and 2019 are compared with the number of observed events seen in each cohort on the panels on the left. Corresponding standardized incidence ratios (SIRs) with 95% confidence intervals (95% CI) are shown in in the panels on the right.

**Table 2.** Observed versus expected events among people vaccinated against SARS-CoV-2 or with a positive PCR test for SARS-CoV-2. For each event of interest, the number of people contributing to the analysis from the target population, their person-years contributed, and the number of observed events are given. Expected events are estimated using indirect standardization to the general population. Standardized incidence ratios (SIRs) with 95% confidence intervals (CIs) were estimated. Events with fewer than 5 occurrences were omitted for privacy reasons: cerebral venous sinus thrombosis and splanchnic vein thrombosis (BNT162b2 and SARS-CoV-2), venous thromboembolism with thrombocytopenia (SARS-CoV-2), and myocardial infarction or ischemic stroke with thrombocytopenia (BNT162b2 and SARS-CoV-2). *Cerebral venous sinus thrombosis is reported without the requirement for a year of prior history as fewer than 5 events were seen when this restriction was imposed.

|  | N | Person-years | Observed events | Expected events | Standardised event difference proportion % | SIR (95% CI) |
| --- | --- | --- | --- | --- | --- | --- |
| **Thrombosis** |  |  |  |  |  |  |
| ***Cerebral venous sinus thrombosis (CVST)\**** |  |  |  |  |  |  |
| Vaccinated with ChAdOx1 | 1,956,136 | 124,569 | 5 | 2.2 | 0.0001% | 2.32 (0.97 to 5.58) |
| ***Deep vein thrombosis (DVT)*** |  |  |  |  |  |  |
| Vaccinated with ChAdOx1 | 1,863,668 | 118,809 | 226 | 237.9 | -0.0006% | 0.95 (0.83 to 1.08) |
| Vaccinated with BNT162b2 | 1,656,875 | 118,344 | 254 | 251.3 | 0.0002% | 1.01 (0.89 to 1.14) |
| SARS-CoV-2 PCR positive test | 298,776 | 40,528 | 125 | 48.8 | 0.0255% | 2.56 (2.15 to 3.05) |
| ***Pulmonary embolism (PE)*** |  |  |  |  |  |  |
| Vaccinated with ChAdOx1 | 1,863,403 | 118,787 | 259 | 210.4 | 0.0026% | 1.23 (1.09 to 1.39) |
| Vaccinated with BNT162b2 | 1,656,755 | 118,336 | 261 | 216.4 | 0.0027% | 1.21 (1.07 to 1.36) |
| SARS-CoV-2 PCR positive test | 298,786 | 40,491 | 547 | 35.7 | 0.1711% | 15.31 (14.08 to 16.65) |
| ***Splanchnic Vein Thrombosis (SVT)*** |  |  |  |  |  |  |
| Vaccinated with ChAdOx1 | 1,868,536 | 119,132 | 9 | 6.5 | 0.0001% | 1.38 (0.72 to 2.66) |
| ***Venous thromboembolism (VTE)*** |  |  |  |  |  |  |
| Vaccinated with ChAdOx1 | 1,858,837 | 118,483 | 461 | 429.2 | 0.0017% | 1.07 (0.98 to 1.18) |
| Vaccinated with BNT162b2 | 1,652,892 | 118,054 | 491 | 448.6 | 0.0026% | 1.09 (1.00 to 1.20) |
| SARS-CoV-2 PCR positive test | 298,397 | 40,429 | 646 | 80 | 0.1897% | 8.08 (7.48 to 8.72) |
| ***Myocardial infarction*** |  |  |  |  |  |  |
| Vaccinated with ChAdOx1 | 1,861,365 | 118,643 | 347 | 411.3 | -0.0035% | 0.84 (0.76 to 0.94) |
| Vaccinated with BNT162b2 | 1,654,037 | 118,145 | 356 | 433.4 | -0.0047% | 0.82 (0.74 to 0.91) |
| SARS-CoV-2 PCR positive test | 298,632 | 40,509 | 79 | 65.7 | 0.0045% | 1.20 (0.96 to 1.50) |
| ***Ischemic stroke*** |  |  |  |  |  |  |
| Vaccinated with ChAdOx1 | 1,867,000 | 119,025 | 72 | 95.8 | -0.0013% | 0.75 (0.60 to 0.95) |
| Vaccinated with BNT162b2 | 1,659,635 | 118,542 | 120 | 108.5 | 0.0007% | 1.11 (0.92 to 1.32) |
| SARS-CoV-2 PCR positive test | 299,100 | 40,582 | 13 | 12.3 | 0.0002% | 1.05 (0.61 to 1.81) |
| ***ATE (Myocardial infarction or ischemic stroke)*** |  |  |  |  |  |  |
| Vaccinated with ChAdOx1 | 1,858,974 | 118,481 | 406 | 503.7 | -0.0053% | 0.81 (0.73 to 0.89) |
| Vaccinated with BNT162b2 | 1,651,903 | 117,990 | 458 | 538.3 | -0.0049% | 0.85 (0.78 to 0.93) |
| SARS-CoV-2 PCR positive test | 298,464 | 40,485 | 88 | 77.6 | 0.0035% | 1.13 (0.92 to 1.40) |
| ***Stroke*** |  |  |  |  |  |  |
| Vaccinated with ChAdOx1 | 1,861,358 | 118,629 | 413 | 405 | 0.0004% | 1.02 (0.93 to 1.12) |
| Vaccinated with BNT162b2 | 1,654,569 | 118,167 | 457 | 454 | 0.0002% | 1.01 (0.92 to 1.10) |
| SARS-CoV-2 PCR positive test | 298,709 | 40,525 | 65 | 54.5 | 0.0035% | 1.19 (0.93 to 1.52) |
| **Thrombocytopenia** |  |  |  |  |  |  |
| *Immune thrombocytopenia* |  |  |  |  |  |  |
| Vaccinated with ChAdOx1 | 1,868,376 | 119,120 | 18 | 8.9 | 0.0005% | 2.01 (1.27 to 3.19) |
| Vaccinated with BNT162b2 | 1,660,786 | 118,628 | 15 | 8.6 | 0.0004% | 1.74 (1.05 to 2.89) |
| SARS-CoV-2 PCR positive test | 299,175 | 40,593 | 5 | 1.8 | 0.0011% | 2.83 (1.18 to 6.80) |
| **Thrombocytopenia** |  |  |  |  |  |  |
| Vaccinated with ChAdOx1 | 1,840,495 | 117,121 | 1,707 | 1,368.00 | 0.0184% | 1.25 (1.19 to 1.31) |
| Vaccinated with BNT162b2 | 1,632,328 | 116,491 | 1,462 | 1,477.10 | -0.0009% | 0.99 (0.94 to 1.04) |
| SARS-CoV-2 PCR positive test | 296,999 | 40,289 | 254 | 248.6 | 0.0018% | 1.02 (0.90 to 1.16) |
| ***VTE with thrombocytopenia*** |  |  |  |  |  |  |
| Vaccinated with ChAdOx1 | 1,868,547 | 119,132 | 7 | 6.4 | 0.0000% | 1.09 (0.52 to 2.29) |
| Vaccinated with BNT162b2 | 1,660,964 | 118,641 | 6 | 6.9 | -0.0001% | 0.86 (0.39 to 1.92) |
| ***ATE with thrombocytopenia*** |  |  |  |  |  |  |
| Vaccinated with ChAdOx1 | 1,868,580 | 119,135 | 5 | 3.2 | 0.0001% | 1.57 (0.65 to 3.78) |
| ***Stroke with thrombocytopenia*** |  |  |  |  |  |  |
| Vaccinated with ChAdOx1 | 1,868,605 | 119,136 | 6 | 2.7 | 0.0002% | 2.21 (0.99 to 4.91) |

Although crude rates of DVT post-vaccination were higher than expected in some strata (Figure 2), standardization attenuated this difference, with SIRs of 0.95 [0.83-1.08] for ChadOx1 and 1.01 [0.89-1.14] for BNT162b2 (Figure 3). DVT rates after SARS-Cov-2 infection were more than double the expected rates, with a SIR of 2.56 [2.15-3.05].

Post-vaccination rates of PE were higher than expected in some strata, such as women and men aged 50-59, women aged >=80 vaccinated with ChAdOx1, and men aged 60-69 vaccinated with BNT162b2 (Figure 2). After standardization, excess PE events remained for ChAdOx1 and Bnt162n2, with SIRs of 1.23 [1.09-1.39] and 1.21 [1.07-1.36], respectively. In comparison, SARS-Cov-2 infection had a SIR of 15.31 [14.08-16.65]. Incidence rate ratios by age and sex for PE and the profiles of those with an event are summarized in the Appendix.

Few occurrences of SVT were seen, with a SIR of 1.38 [0.72-2.66] for ChAdOx1 and under 5 cases following BNT162b2 vaccination or SARS-Cov-2 infection. CVST was too rare (n<5) in all three cohorts to conduct the primary analyses (see Figure 3 and below for results from sensitivity analyses).

Observed post-vaccination ATE rates were similar to expected rates (Figure 2). There were fewer ATE than expected for both vaccinated cohorts after standardization (Table 2, Figure 3). Little differences was seen between observed and expected ATE in people infected with SARS-Cov-2 (Figure 3). When analyzed separately, observed rates of myocardial infarction and ischemic stroke were also similar to expected rates following both vaccination and infection with SARS-Cov-2 (Figure 2, Figure 3).

Thrombocytopenia was more common than expected after vaccination with ChAdOx1 (SIR 1.25 [1.19-1.31]), but not after vaccination with BNT162b2 (SIR 0.99 [0.94-1.04]). Immune thrombocytopenia was more common than expected from the background population after vaccination with ChAdOx1 (SIR 2.01 [1.27-3.19]), BNT162b2 (SIR 1.74 [1.05-2.89]), and SARS-Cov-2 infection (SIR 2.83 [1.18-6.80]). Incidence rate ratios by age and sex for thrombocytopenia are summarized in the Appendix, along with the profiles of those affected.

VTE with concurrent thrombocytopenia was very rare and had similar crude rates in the background population (3.5/100,000 person-years [3.1-4.1]), after ChAdOx1 vaccination (5.9 [2.4-12.1]), and after BNT162b2 vaccination (5.1 [1.9-11.0]). There were fewer than 5 cases in the SARS-Cov-2 cohort. Standardization confirmed this finding, with SIRs of 1.09 [0.52- 2.29] for ChAdOx1 and 0.86 [0.39-1.92] for BNT162b2.

ATE with thrombocytopenia was also very rare, with a rate after ChAdOx1 vaccination of 4.2/100,000 person-years [1.4-9.8] and an overall crude background rate of 1.7/100,000 person-years [1.4-2.1], equivalent to a SIR of 1.57 [0.65-3.78]. Fewer than 5 cases were identified after BNT162b2 vaccination and SARS-Cov-2 infection.

Overall crude rates of stroke with thrombocytopenia were 1.4/100,000 person-years [1.1-1.8] in the background population and 5.0 [1.8-11.0] after vaccination with ChAdOx1, equivalent to a SIR of 2.21 [0.99 to 4.91]. Again, under 5 cases were seen after BNT162b2 vaccination and SARS-Cov-2 infection, so SIR rates were not estimated. Myocardial infarction with thrombocytopenia was too rare (n<5) in the vaccination and infection cohorts for analysis.

### Sensitivity analyses

Results from sensitivity analyses were generally consistent with the results from the primary analyses (see Appendix). One key addition was the analysis of CVST, which was too rare for estimation in the primary analyses but could be analyzed when the requirement of a year of prior history was removed. The observed number of CVST events (n=5) was then higher than expected (2.2) in the ChAdOx1 cohort, resulting in a SIR of 2.32 [0.97-5.58] (Table 2, Figure 3).

All primary and sensitivity analysis results are available in an interactive web application: https://livedataoxford.shinyapps.io/CovidVaccinationSafetyStudy/

## Discussion

### Summary of results

In a cohort of 3.5 million people vaccinated against SARS-CoV-2, 74 (0.002%) more than expected suffered a VTE in the 28 days following vaccination. This higher rate was driven by more PE events after both ChAdOx1 and BNT162b2 than expected, with a 20% increase in rates of events and 93 (0.003%) excess PEs observed. The rate of PE was much larger in people infected with SARS-Cov-2, with a 15-fold relative increase and 511 (0.17%) excess PE events observed in 300,000 people.

Although observed rates of thrombocytopenia were similar to expected for people vaccinated with BNT162b2, a 25% relative increase in the rate of thrombocytopenia was seen among those vaccinated with ChAdOx1. This increase was most pronounced for younger age groups. No signal was seen for either vaccine for ATE. Finally, 3 (0.0001%) more cases than expected were seen of CVST and 3 (0.0002%) of stroke with thrombocytopenia in 1.8 million people vaccinated with ChAdOx1 in a secondary analysis.

Noticeable differences in comorbidities between the vaccinated and background cohorts suggest the potential for residual confounding, unaccounted for in our analyses.

### Findings in context

Concerns over thrombosis – alone and with thrombocytopenia – have been raised from spontaneous reports data since March.^8, 9, 20^ Case series have been published, suggesting a new clinical entity known as vaccine-induced immune thrombotic thrombocytopenia (VITT), presenting as unusual thrombosis with raised antibodies against platelet factor 4. Only a few small studies have been conducted on this topic. A study of 280,000 vaccinees aged between 18 and 65 in Denmark and Norway assessed the 28-day incidence rates of thromboembolic events and coagulation disorders following ChAdOx1.^21^ Similar to our analyses, Pottegård et al applied a historical comparator design with indirect standardization. They found a 2-fold increased rate of VTE, an 80% increase in rates of PE, and a 20-fold increased rate of CVST. The authors also reported a 3-fold higher-than-expected rate of thrombocytopenia and a potential increase. As in our study, they observed similar rates of arterial events among those vaccinated as would be expected given rates in the general population.

More recently, a nested case-control study from Scotland found no increase in risk of VTE with either vaccine.^22^ The authors also reported potential increased risks of ATE and hemorrhagic events with ChAdOx1, although these were not confirmed in subsequent self-controlled case series analysis. The authors suggested that residual confounding could explain their findings. Case-control analyses based on routine health data have recently been criticized and shown to induce substantial residual bias,^23^ which may explain the differences in results from those in our study and that by Pottegård et al.

### Study strengths and limitations

Our study has limitations. The time period studied covered the initial phases of vaccination in the UK, when vaccines were prioritized for older, more vulnerable populations and healthcare staff.^24^ We therefore saw a higher prevalence of conditions such as asthma and diabetes in those vaccinated than in the general population. Although we used indirect standardization to account for differences in the four cohorts’ age distributions, remaining residual confounding could explain some of our findings. Such bias could result in overestimated safety signals due to remaining imbalances in the baseline outcome risk when comparing vaccinated and background populations.

Measurement error is unavoidable in observational studies. However, any errors are likely to have been non-differential across our vaccinated and unvaccinated cohorts and should therefore not have affected our relative rate estimates. As we only used primary care data, we may have underestimated absolute risks due to a lack of hospital linkage. However, previous studies have shown that CPRD captures rare events well, even without linkage to Hospital Episode Statistics.^25^

Although within-database comparisons are preferred,^16^ we had to use two slightly different primary care databases. However, one study found that these databases gave similar results.^26^ We also used the OMOP common data model to maximize the comparability of the two databases irrespective of differences in their coding systems.

Our study also has strengths. This is the largest cohort study on the safety of COVID-19 vaccines to date. The large sample of 3.5 million vaccinees allowed us to assess very rare events that are generally not observed in clinical trials. We used a well-established source of routinely collected health data previously used for vaccine safety studies.^27, 28^ Including cohorts of people infected with SARS-CoV-2 and the general population provided much needed context for interpreting our findings. Our historical comparison method ensured a timely study with high statistical power. Our protocol was pre-specified and agreed in advance with the study funder, the European Medicines Agency. Our analyses were based on data recorded before the detection of thrombosis and thrombocytopenia signals, minimizing surveillance bias.

## Conclusions

In a cohort of 3.5 million people vaccinated against SARS-Cov-2, thrombosis, thrombocytopenia, and thrombosis with thrombocytopenia were very rare events. The rates of VTE and ATE were broadly similar after ChAdOx1 and BNT162b2 vaccination, although thrombocytopenia was more common among ChAdOx1 vaccinees, particularly in younger age strata. A potential safety signal for PE for both vaccines was identified. Although the occurrence of PE after vaccination was 1.2-fold above that expected in the general population, its occurrence among those contracting SARS-CoV-2 was more than 15-fold the background (expected) rate. Vaccinated people recorded more comorbidities and medicine use than the background population, suggesting potential for unresolved confounding by indication.

## Data Availability

In accordance with current European and national law, the data used in this study is only available for the researchers participating in this study. Thus, we are not allowed to distribute or make publicly available the data to other parties. However, researchers from public institutions can request data from SIDIAP if they comply with certain requirements. Further information is available online (https://www.sidiap.org/index.php/menu-solicitudes-en/application-proccedure) or by contacting Anna Moleras.

## Ethical approvals

The protocol for this research was approved by the Independent Scientific Advisory Committee (ISAC) for MHRA Database Research (protocol number 20_000211).

## Acknowledgements

This study was funded by the European Medicines Agency in the form of a competitive tender (Lot ROC No EMA/2017/09/PE). We acknowledge Prof Johan Van der Lei for the overall management of this research grant.

## Appendix

### Patient characteristics by age group

#### Characteristics of study participants aged: 30 to 44

The characteristics of the study cohorts used for the primary analyses, all with the requirement to be aged 30 years or older and with a year of prior history observed in the database. Those in the general population had a primary care visit/ contact between 2017 and 2019 while persons infected with SARS-CoV-2 had a confirmatory positive RT-PCR test. *Conditions of interest: autoimmune disease, antiphospholipid syndrome, thrombophilia, asthma, atrial fibrillation, malignant neoplastic disease, diabetes mellitus, obesity, or renal impairment. ^†^Medications of interest included non-steroidal anti-inflammatory drugs, Cox2 inhibitors, systemic corticosteroids, hormonal contraceptives, tamoxifen, and sex hormones and modulators of the genital system

|  | General population | Vaccinated with ChAdOx1 | Vaccinated with BNT162b2 | SARS-CoV-2 PCR positive test |
| --- | --- | --- | --- | --- |
| N | 644,372 | 202,865 | 217,319 | 121,692 |
| Age | 37 [33 to 41] | 38 [34 to 41] | 38 [34 to 41] | 37 [33 to 41] |
| Age: 30 to 39 | 431,801 (67.0%) | 123,991 (61.1%) | 135,101 (62.2%) | 82,427 (67.7%) |
| Age: 40 to 49 | 212,571 (33.0%) | 78,874 (38.9%) | 82,218 (37.8%) | 39,265 (32.3%) |
| Sex: Male | 306,045 (47.5%) | 74,116 (36.5%) | 70,735 (32.5%) | 53,806 (44.2%) |
| Years of prior observation time | 10.4 [4.8 to 14.9] | 8.3 [4.1 to 15.9] | 7.7 [3.8 to 14.9] | 7.9 [3.8 to 15.7] |
| Comorbidities |  |  |  |  |
| Autoimmune disease | 8,875 (1.4%) | 5,779 (2.8%) | 5,283 (2.4%) | 1,847 (1.5%) |
| Antiphospholipid syndrome | 259 (0.0%) | 273 (0.1%) | 251 (0.1%) | 76 (0.1%) |
| Thrombophilia | 851 (0.1%) | 660 (0.3%) | 654 (0.3%) | 227 (0.2%) |
| Asthma | 102,513 (15.9%) | 47,467 (23.4%) | 46,382 (21.3%) | 20,043 (16.5%) |
| Atrial fibrillation | 927 (0.1%) | 879 (0.4%) | 863 (0.4%) | 192 (0.2%) |
| Malignant neoplastic disease | 7,288 (1.1%) | 5,844 (2.9%) | 5,492 (2.5%) | 1,356 (1.1%) |
| Diabetes mellitus | 17,185 (2.7%) | 27,531 (13.6%) | 19,860 (9.1%) | 5,576 (4.6%) |
| Obesity | 18,089 (2.8%) | 13,635 (6.7%) | 11,684 (5.4%) | 4,815 (4.0%) |
| Heart disease | 11,653 (1.8%) | 10,509 (5.2%) | 9,827 (4.5%) | 2,886 (2.4%) |
| Hypertensive disorder | 20,630 (3.2%) | 13,490 (6.6%) | 12,430 (5.7%) | 4,910 (4.0%) |
| Renal impairment | 4,449 (0.7%) | 4,107 (2.0%) | 3,695 (1.7%) | 1,149 (0.9%) |
| COPD | 929 (0.1%) | 911 (0.4%) | 613 (0.3%) | 96 (0.1%) |
| Dementia | 772 (0.1%) | 738 (0.4%) | 638 (0.3%) | 206 (0.2%) |
| Medication use (183 days prior to four days prior) |  |  |  |  |
| Non-steroidal anti-inflammatory drugs | 107,115 (16.6%) | 20,685 (10.2%) | 18,571 (8.5%) | 9,204 (7.6%) |
| Cox2 inhibitors | 933 (0.1%) | 183 (0.1%) | 147 (0.1%) | 54 (0.0%) |
| Systemic corticosteroids | 44,535 (6.9%) | 10,629 (5.2%) | 9,351 (4.3%) | 4,406 (3.6%) |
| Antithrombotic and anticoagulant therapies | 4,599 (0.7%) | 1,031 (0.5%) | 789 (0.4%) | 269 (0.2%) |
| Lipid modifying agents | 4,098 (0.6%) | 1,861 (0.9%) | 1,543 (0.7%) | 490 (0.4%) |
| Antineoplastic and immunomodulating agents | 29,367 (4.6%) | 5,402 (2.7%) | 6,002 (2.8%) | 2,591 (2.1%) |
| Hormonal contraceptives for systemic use | 50,360 (7.8%) | 9,897 (4.9%) | 11,305 (5.2%) | 5,000 (4.1%) |
| Tamoxifen | 190 (0.0%) | 47 (0.0%) | 44 (0.0%) | 14 (0.0%) |
| Sex hormones and modulators of the genital system | 52,721 (8.2%) | 10,572 (5.2%) | 11,987 (5.5%) | 5,268 (4.3%) |
| One or more condition of interest* | 52,792 (8.2%) | 50,913 (25.1%) | 41,923 (19.3%) | 13,637 (11.2%) |
| One or more medication of interest<sup>†</sup> | 158,520 (24.6%) | 33,547 (16.5%) | 32,307 (14.9%) | 15,456 (12.7%) |
| One or more condition/ medication of interest*<sup>†</sup> | 191,069 (29.7%) | 73,997 (36.5%) | 65,853 (30.3%) | 26,342 (21.6%) |

#### Characteristics of study participants: aged 45 to 64

The characteristics of the study cohorts used for the primary analyses, all with the requirement to be aged 30 years or older and with a year of prior history observed in the database. Those in the general population had a primary care visit/ contact between 2017 and 2019 while persons infected with SARS-CoV-2 had a confirmatory positive RT-PCR test. *Conditions of interest: autoimmune disease, antiphospholipid syndrome, thrombophilia, asthma, atrial fibrillation, malignant neoplastic disease, diabetes mellitus, obesity, or renal impairment. ^†^Medications of interest included non-steroidal anti-inflammatory drugs, Cox2 inhibitors, systemic corticosteroids, hormonal contraceptives, tamoxifen, and sex hormones and modulators of the genital system

|  | General population | Vaccinated with ChAdOx1 | Vaccinated with BNT162b2 | SARS-CoV-2 PCR positive test |
| --- | --- | --- | --- | --- |
| N | 963,696 | 624,698 | 534,049 | 138,195 |
| Age | 54 [49 to 59] | 57 [52 to 62] | 56 [51 to 60] | 53 [49 to 58] |
| Age: 40 to 49 | 246,829 (25.6%) | 94,579 (15.1%) | 96,285 (18.0%) | 37,919 (27.4%) |
| Age: 50 to 59 | 506,515 (52.6%) | 285,938 (45.8%) | 279,223 (52.3%) | 75,543 (54.7%) |
| Age: 60 to 69 | 210,352 (21.8%) | 244,181 (39.1%) | 158,541 (29.7%) | 24,733 (17.9%) |
| Age: 70 to 79 | 0 (0.0%) | 0 (0.0%) | 0 (0.0%) | 0 (0.0%) |
| Age: 80 or older | 0 (0.0%) | 0 (0.0%) | 0 (0.0%) | 0 (0.0%) |
| Sex: Male | 476,832 (49.5%) | 287,869 (46.1%) | 215,350 (40.3%) | 64,807 (46.9%) |
| Years of prior observation time | 13.8 [9.4 to 16.5] | 16.9 [7.5 to 27.0] | 16.6 [7.6 to 26.4] | 16.0 [7.4 to 25.3] |
| Comorbidities |  |  |  |  |
| Autoimmune disease | 22,807 (2.4%) | 24,114 (3.9%) | 20,452 (3.8%) | 3,497 (2.5%) |
| Antiphospholipid syndrome | 553 (0.1%) | 785 (0.1%) | 670 (0.1%) | 125 (0.1%) |
| Thrombophilia | 1,247 (0.1%) | 1,721 (0.3%) | 1,511 (0.3%) | 284 (0.2%) |
| Asthma | 120,450 (12.5%) | 107,823 (17.3%) | 94,363 (17.7%) | 19,830 (14.3%) |
| Atrial fibrillation | 10,190 (1.1%) | 12,981 (2.1%) | 12,425 (2.3%) | 1,594 (1.2%) |
| Malignant neoplastic disease | 49,036 (5.1%) | 53,877 (8.6%) | 47,018 (8.8%) | 6,929 (5.0%) |
| Diabetes mellitus | 70,192 (7.3%) | 102,585 (16.4%) | 90,441 (16.9%) | 14,163 (10.2%) |
| Obesity | 43,294 (4.5%) | 48,229 (7.7%) | 40,960 (7.7%) | 9,226 (6.7%) |
| Heart disease | 60,885 (6.3%) | 73,169 (11.7%) | 66,530 (12.5%) | 9,890 (7.2%) |
| Hypertensive disorder | 173,364 (18.0%) | 164,162 (26.3%) | 137,388 (25.7%) | 28,262 (20.5%) |
| Renal impairment | 26,427 (2.7%) | 32,833 (5.3%) | 28,361 (5.3%) | 4,445 (3.2%) |
| COPD | 21,130 (2.2%) | 27,484 (4.4%) | 20,039 (3.8%) | 2,056 (1.5%) |
| Dementia | 1,914 (0.2%) | 2,642 (0.4%) | 1,983 (0.4%) | 370 (0.3%) |
| Medication use (183 days prior to four days prior) |  |  |  |  |
| Non-steroidal anti-inflammatory drugs | 234,513 (24.3%) | 79,342 (12.7%) | 65,776 (12.3%) | 15,556 (11.3%) |
| Cox2 inhibitors | 3,024 (0.3%) | 793 (0.1%) | 609 (0.1%) | 161 (0.1%) |
| Systemic corticosteroids | 97,185 (10.1%) | 36,413 (5.8%) | 29,039 (5.4%) | 6,364 (4.6%) |
| Antithrombotic and anticoagulant therapies | 25,233 (2.6%) | 12,124 (1.9%) | 10,842 (2.0%) | 1,489 (1.1%) |
| Lipid modifying agents | 47,616 (4.9%) | 28,586 (4.6%) | 24,220 (4.5%) | 4,436 (3.2%) |
| Antineoplastic and immunomodulating agents | 14,370 (1.5%) | 5,635 (0.9%) | 4,958 (0.9%) | 874 (0.6%) |
| Hormonal contraceptives for systemic use | 25,338 (2.6%) | 7,644 (1.2%) | 8,444 (1.6%) | 2,172 (1.6%) |
| Tamoxifen | 1,095 (0.1%) | 449 (0.1%) | 438 (0.1%) | 61 (0.0%) |
| Sex hormones and modulators of the genital system | 43,992 (4.6%) | 19,408 (3.1%) | 20,440 (3.8%) | 4,542 (3.3%) |
| One or more condition of interest* | 187,548 (19.5%) | 221,112 (35.4%) | 195,178 (36.5%) | 33,104 (24.0%) |
| One or more medication of interest† | 288,508 (29.9%) | 108,882 (17.4%) | 93,553 (17.5%) | 21,648 (15.7%) |
| One or more condition/medication of interest*† | 395,455 (41.0%) | 283,344 (45.4%) | 248,153 (46.5%) | 47,564 (34.4%) |

#### Characteristics of study participants: age 65 or older

The characteristics of the study cohorts used for the primary analyses, all with the requirement to be aged 30 years or older and with a year of prior history observed in the database. Those in the general population had a primary care visit/ contact between 2017 and 2019 while persons infected with SARS-CoV-2 had a confirmatory positive RT-PCR test. *Conditions of interest: autoimmune disease, antiphospholipid syndrome, thrombophilia, asthma, atrial fibrillation, malignant neoplastic disease, diabetes mellitus, obesity, or renal impairment. ^†^Medications of interest included non-steroidal anti-inflammatory drugs, Cox2 inhibitors, systemic corticosteroids, hormonal contraceptives, tamoxifen, and sex hormones and modulators of the genital system

|  | General population | Vaccinated with ChAdOx1 | Vaccinated with BNT162b2 | SARS-CoV-2 PCR positive test |
| --- | --- | --- | --- | --- |
| N | 682,469 | 1,041,204 | 909,771 | 39,424 |
| Age | 74 [69 to 80] | 73 [69 to 77] | 77 [71 to 83] | 73 [68 to 80] |
| Age: 60 to 69 | 193,321 (28.3%) | 322,860 (31.0%) | 163,711 (18.0%) | 13,913 (35.3%) |
| Age: 70 to 79 | 301,179 (44.1%) | 545,251 (52.4%) | 372,426 (40.9%) | 14,839 (37.6%) |
| Age: 80 or older | 187,969 (27.5%) | 173,093 (16.6%) | 373,634 (41.1%) | 10,672 (27.1%) |
| Sex: Male | 312,034 (45.7%) | 483,923 (46.5%) | 416,778 (45.8%) | 17,688 (44.9%) |
| Years of prior observation time | 14.3 [12.2 to 17.3] | 23.6 [10.1 to 34.6] | 24.9 [11.4 to 35.6] | 20.6 [6.8 to 32.8] |
| Comorbidities |  |  |  |  |
| Autoimmune disease | 26,091 (3.8%) | 42,226 (4.1%) | 38,576 (4.2%) | 1,792 (4.5%) |
| Antiphospholipid syndrome | 234 (0.0%) | 477 (0.0%) | 428 (0.0%) | 14 (0.0%) |
| Thrombophilia | 484 (0.1%) | 1,247 (0.1%) | 1,027 (0.1%) | 38 (0.1%) |
| Asthma | 78,706 (11.5%) | 128,836 (12.4%) | 112,432 (12.4%) | 5,597 (14.2%) |
| Atrial fibrillation | 61,526 (9.0%) | 86,870 (8.3%) | 104,110 (11.4%) | 4,207 (10.7%) |
| Malignant neoplastic disease | 131,175 (19.2%) | 197,728 (19.0%) | 216,145 (23.8%) | 7,378 (18.7%) |
| Diabetes mellitus | 110,242 (16.2%) | 176,828 (17.0%) | 165,893 (18.2%) | 8,749 (22.2%) |
| Obesity | 32,671 (4.8%) | 58,188 (5.6%) | 46,256 (5.1%) | 2,750 (7.0%) |
| Heart disease | 179,170 (26.3%) | 250,791 (24.1%) | 273,312 (30.0%) | 11,735 (29.8%) |
| Hypertensive disorder | 339,756 (49.8%) | 500,091 (48.0%) | 493,527 (54.2%) | 20,749 (52.6%) |
| Renal impairment | 130,720 (19.2%) | 170,811 (16.4%) | 202,330 (22.2%) | 8,556 (21.7%) |
| COPD | 55,146 (8.1%) | 79,241 (7.6%) | 75,002 (8.2%) | 3,232 (8.2%) |
| Dementia | 26,690 (3.9%) | 35,209 (3.4%) | 32,208 (3.5%) | 4,310 (10.9%) |
| Medication use (183 days prior to four days prior) |  |  |  |  |
| Non-steroidal anti-inflammatory drugs | 276,924 (40.6%) | 164,063 (15.8%) | 157,962 (17.4%) | 8,539 (21.7%) |
| Cox2 inhibitors | 2,158 (0.3%) | 955 (0.1%) | 825 (0.1%) | 43 (0.1%) |
| Systemic corticosteroids | 121,768 (17.8%) | 72,724 (7.0%) | 67,587 (7.4%) | 3,398 (8.6%) |
| Antithrombotic and anticoagulant therapies | 72,773 (10.7%) | 48,167 (4.6%) | 53,606 (5.9%) | 2,127 (5.4%) |
| Lipid modifying agents | 78,550 (11.5%) | 79,882 (7.7%) | 77,072 (8.5%) | 3,298 (8.4%) |
| Antineoplastic and immunomodulating agents | 13,666 (2.0%) | 9,408 (0.9%) | 9,581 (1.1%) | 357 (0.9%) |
| Hormonal contraceptives for systemic use | 1,098 (0.2%) | 1,025 (0.1%) | 742 (0.1%) | 33 (0.1%) |
| Tamoxifen | 672 (0.1%) | 318 (0.0%) | 317 (0.0%) | 12 (0.0%) |
| Sex hormones and modulators of the genital system | 13,870 (2.0%) | 14,632 (1.4%) | 12,136 (1.3%) | 473 (1.2%) |
| One or more condition of interest* | 339,288 (49.7%) | 506,534 (48.6%) | 514,123 (56.5%) | 22,138 (56.2%) |
| One or more medication of interest<sup>†</sup> | 311,320 (45.6%) | 203,446 (19.5%) | 192,844 (21.2%) | 10,135 (25.7%) |
| One or more condition/ medication of interest*<sup>†</sup> | 471,617 (69.1%) | 591,852 (56.8%) | 583,751 (64.2%) | 25,821 (65.5%) |

### Results for all study outcomes

For each event of interest the number of persons contributing to the analysis from the target population, their person-years contributed, and the number of events observed for them are given. Their expected events are estimated using indirect standardisation to the general population, with expected events giving the number of events we would have expected to have seen if their outcome experience was the same as that of the general population. Standardised incidence ratios (SIRs) with 95% confidence intervals (CIs) were estimated. Events with less than 5 occurrences have been omitted for privacy reasons. *Cerebral venous sinus thrombosis is reported without the requirement for a year of prior history as less than 5 events were seen when this restriction was imposed.

| Event | Cohort | N | Person-years | Observed events | Expected events | SIR (95% CI) |
| --- | --- | --- | --- | --- | --- | --- |
| Cerebral venous sinus thrombosis* | Vaccinated with ChAdOx1 | 1,956,136 | 124,569 | 5 | 2.2 | 2.32 (0.97 to 5.58) |
| Deep vein thrombosis - broad | Vaccinated with ChAdOx1 | 1,862,896 | 118,757 | 275 | 255.1 | 1.08 (0.96 to 1.21) |
| Deep vein thrombosis - broad | Vaccinated with BNT162b2 | 1,656,235 | 118,297 | 296 | 268.1 | 1.10 (0.99 to 1.24) |
| Deep vein thrombosis - broad | SARS-CoV-2 PCR positive test | 298,707 | 40,517 | 143 | 53.2 | 2.69 (2.28 to 3.17) |
| Deep vein thrombosis - narrow | Vaccinated with ChAdOx1 | 1,863,668 | 118,809 | 226 | 237.9 | 0.95 (0.83 to 1.08) |
| Deep vein thrombosis - narrow | Vaccinated with BNT162b2 | 1,656,875 | 118,344 | 254 | 251.3 | 1.01 (0.89 to 1.14) |
| Deep vein thrombosis - narrow | SARS-CoV-2 PCR positive test | 298,776 | 40,528 | 125 | 48.8 | 2.56 (2.15 to 3.05) |
| Deep vein thrombosis broad with thrombocytopenia 10 days pre to 10 days post | Vaccinated with ChAdOx1 | 1,868,584 | 119,135 | 6 | 4.1 | 1.45 (0.65 to 3.23) |
| Deep vein thrombosis broad with thrombocytopenia 42 days pre to 14 days post | Vaccinated with ChAdOx1 | 1,868,547 | 119,132 | 9 | 7.4 | 1.22 (0.64 to 2.35) |
| Deep vein thrombosis narrow with thrombocytopenia 42 days pre to 14 days post | Vaccinated with ChAdOx1 | 1,868,554 | 119,133 | 7 | 7 | 1.00 (0.48 to 2.11) |
| Hemorrhagic stroke | Vaccinated with ChAdOx1 | 1,867,822 | 119,084 | 45 | 43.1 | 1.04 (0.78 to 1.40) |
| Hemorrhagic stroke | Vaccinated with BNT162b2 | 1,660,431 | 118,603 | 38 | 46.8 | 0.81 (0.59 to 1.12) |
| Hemorrhagic stroke | SARS-CoV-2 PCR positive test | 299,127 | 40,584 | 11 | 8.2 | 1.34 (0.74 to 2.42) |
| Immune thrombocytopenia | Vaccinated with ChAdOx1 | 1,868,376 | 119,120 | 18 | 8.9 | 2.01 (1.27 to 3.19) |
| Immune thrombocytopenia | Vaccinated with BNT162b2 | 1,660,786 | 118,628 | 15 | 8.6 | 1.74 (1.05 to 2.89) |
| Immune thrombocytopenia | SARS-CoV-2 PCR positive test | 299,175 | 40,593 | 5 | 1.8 | 2.83 (1.18 to 6.80) |
| Intestinal infarction | Vaccinated with ChAdOx1 | 1,868,365 | 119,120 | 13 | 15.1 | 0.86 (0.50 to 1.48) |
| Intestinal infarction | Vaccinated with BNT162b2 | 1,660,825 | 118,631 | 17 | 15.5 | 1.10 (0.68 to 1.77) |
| Ischemic stroke | Vaccinated with ChAdOx1 | 1,867,000 | 119,025 | 72 | 95.8 | 0.75 (0.60 to 0.95) |
| Ischemic stroke | Vaccinated with BNT162b2 | 1,659,635 | 118,542 | 120 | 108.5 | 1.11 (0.92 to 1.32) |
| Ischemic stroke | SARS-CoV-2 PCR positive test | 299,100 | 40,582 | 13 | 12.3 | 1.05 (0.61 to 1.81) |
| Myocardial infarction | Vaccinated with ChAdOx1 | 1,861,365 | 118,643 | 347 | 411.3 | 0.84 (0.76 to 0.94) |
| Myocardial infarction | Vaccinated with BNT162b2 | 1,654,037 | 118,145 | 356 | 433.4 | 0.82 (0.74 to 0.91) |
| Myocardial infarction | SARS-CoV-2 PCR positive test | 298,632 | 40,509 | 79 | 65.7 | 1.20 (0.96 to 1.50) |
| Myocardial infarction or ischemic stroke | Vaccinated with ChAdOx1 | 1,858,974 | 118,481 | 406 | 503.7 | 0.81 (0.73 to 0.89) |
| Myocardial infarction or ischemic stroke | Vaccinated with BNT162b2 | 1,651,903 | 117,990 | 458 | 538.3 | 0.85 (0.78 to 0.93) |
| Myocardial infarction or ischemic stroke | SARS-CoV-2 PCR positive test | 298,464 | 40,485 | 88 | 77.6 | 1.13 (0.92 to 1.40) |
| Myocardial infarction or ischemic stroke with thrombocytopenia 10 days pre to 10 days post | Vaccinated with ChAdOx1 | 1,868,580 | 119,135 | 5 | 3.2 | 1.57 (0.65 to 3.78) |
| Myocardial infarction or ischemic stroke with thrombocytopenia 42 days pre to 14 days post | Vaccinated with ChAdOx1 | 1,868,524 | 119,131 | 10 | 6.5 | 1.53 (0.82 to 2.84) |
| Myocardial infarction or ischemic stroke with thrombocytopenia 42 days pre to 14 days post | Vaccinated with BNT162b2 | 1,660,931 | 118,639 | 6 | 8.3 | 0.72 (0.32 to 1.61) |
| Platelet disorder | Vaccinated with ChAdOx1 | 1,865,323 | 118,905 | 198 | 86.6 | 2.29 (1.99 to 2.63) |
| Platelet disorder | Vaccinated with BNT162b2 | 1,657,943 | 118,416 | 174 | 88.1 | 1.98 (1.70 to 2.29) |
| Platelet disorder | SARS-CoV-2 PCR positive test | 298,922 | 40,552 | 57 | 15.9 | 3.60 (2.77 to 4.66) |
| Portal vein thrombosis | Vaccinated with ChAdOx1 | 1,868,539 | 119,133 | 8 | 5.4 | 1.48 (0.74 to 2.95) |
| Pulmonary embolism | Vaccinated with ChAdOx1 | 1,863,403 | 118,787 | 259 | 210.4 | 1.23 (1.09 to 1.39) |
| Pulmonary embolism | Vaccinated with BNT162b2 | 1,656,755 | 118,336 | 261 | 216.4 | 1.21 (1.07 to 1.36) |
| Pulmonary embolism | SARS-CoV-2 PCR positive test | 298,786 | 40,491 | 547 | 35.7 | 15.31 (14.08 to 16.65) |
| Pulmonary embolism with thrombocytopenia 42 days pre to 14 days post | Vaccinated with ChAdOx1 | 1,868,576 | 119,134 | 6 | 5.5 | 1.09 (0.49 to 2.43) |
| Pulmonary embolism with thrombocytopenia 42 days pre to 14 days post | Vaccinated with BNT162b2 | 1,660,999 | 118,644 | 7 | 5.9 | 1.18 (0.56 to 2.48) |
| Pulmonary embolism with thrombocytopenia 42 days pre to 14 days post | SARS-CoV-2 PCR positive test | 299,208 | 40,596 | 6 | 0.7 | 8.25 (3.71 to 18.37) |
| Splanchnic Vein Thrombosis | Vaccinated with ChAdOx1 | 1,868,536 | 119,132 | 9 | 6.5 | 1.38 (0.72 to 2.66) |
| Stroke | Vaccinated with ChAdOx1 | 1,861,358 | 118,629 | 413 | 405 | 1.02 (0.93 to 1.12) |
| Stroke | Vaccinated with BNT162b2 | 1,654,569 | 118,167 | 457 | 454 | 1.01 (0.92 to 1.10) |
| Stroke | SARS-CoV-2 PCR positive test | 298,709 | 40,525 | 65 | 54.5 | 1.19 (0.93 to 1.52) |
| Stroke with thrombocytopenia 10 days pre to 10 days post | Vaccinated with ChAdOx1 | 1,868,605 | 119,136 | 6 | 2.7 | 2.21 (0.99 to 4.91) |
| Stroke with thrombocytopenia 42 days pre to 14 days post | Vaccinated with ChAdOx1 | 1,868,573 | 119,134 | 6 | 5.8 | 1.03 (0.46 to 2.29) |
| Stroke with thrombocytopenia 42 days pre to 14 days post | Vaccinated with BNT162b2 | 1,660,982 | 118,643 | 8 | 6.9 | 1.16 (0.58 to 2.32) |
| Thrombocytopenia | Vaccinated with ChAdOx1 | 1,840,495 | 117,121 | 1,707 | 1,368.00 | 1.25 (1.19 to 1.31) |
| Thrombocytopenia | Vaccinated with BNT162b2 | 1,632,328 | 116,491 | 1,462 | 1,477.10 | 0.99 (0.94 to 1.04) |
| Thrombocytopenia | SARS-CoV-2 PCR positive test | 296,999 | 40,289 | 254 | 248.6 | 1.02 (0.90 to 1.16) |
| Thrombocytopenic purpura | Vaccinated with ChAdOx1 | 1,868,562 | 119,134 | 12 | 8 | 1.49 (0.85 to 2.63) |
| Thrombocytopenic purpura | Vaccinated with BNT162b2 | 1,660,955 | 118,641 | 5 | 7.9 | 0.64 (0.26 to 1.53) |
| Venous thromboembolism - broad | Vaccinated with ChAdOx1 | 1,858,064 | 118,431 | 508 | 445.4 | 1.14 (1.05 to 1.24) |
| Venous thromboembolism - broad | Vaccinated with BNT162b2 | 1,652,244 | 118,007 | 532 | 464.6 | 1.15 (1.05 to 1.25) |
| Venous thromboembolism - broad | SARS-CoV-2 PCR positive test | 298,329 | 40,418 | 664 | 84.2 | 7.89 (7.31 to 8.51) |
| Venous thromboembolism - narrow | Vaccinated with ChAdOx1 | 1,858,837 | 118,483 | 461 | 429.2 | 1.07 (0.98 to 1.18) |
| Venous thromboembolism - narrow | Vaccinated with BNT162b2 | 1,652,892 | 118,054 | 491 | 448.6 | 1.09 (1.00 to 1.20) |
| Venous thromboembolism - narrow | SARS-CoV-2 PCR positive test | 298,397 | 40,429 | 646 | 80 | 8.08 (7.48 to 8.72) |
| Venous thromboembolism broad with thrombocytopenia 10 days pre to 10 days post | Vaccinated with ChAdOx1 | 1,868,543 | 119,132 | 9 | 6.7 | 1.35 (0.70 to 2.60) |
| Venous thromboembolism broad with thrombocytopenia 10 days pre to 10 days post | Vaccinated with BNT162b2 | 1,660,959 | 118,641 | 6 | 7.1 | 0.84 (0.38 to 1.87) |
| Venous thromboembolism broad with thrombocytopenia 42 days pre to 14 days post | Vaccinated with ChAdOx1 | 1,868,467 | 119,126 | 14 | 12.3 | 1.13 (0.67 to 1.92) |
| Venous thromboembolism broad with thrombocytopenia 42 days pre to 14 days post | Vaccinated with BNT162b2 | 1,660,896 | 118,636 | 10 | 13.3 | 0.75 (0.41 to 1.40) |
| Venous thromboembolism broad with thrombocytopenia 42 days pre to 14 days post | SARS-CoV-2 PCR positive test | 299,199 | 40,595 | 6 | 1.7 | 3.56 (1.60 to 7.93) |
| Venous thromboembolism narrow with thrombocytopenia 10 days pre to 10 days post | Vaccinated with ChAdOx1 | 1,868,547 | 119,132 | 7 | 6.4 | 1.09 (0.52 to 2.29) |
| Venous thromboembolism narrow with thrombocytopenia 10 days pre to 10 days post | Vaccinated with BNT162b2 | 1,660,964 | 118,641 | 6 | 6.9 | 0.86 (0.39 to 1.92) |
| Venous thromboembolism narrow with thrombocytopenia 42 days pre to 14 days post | Vaccinated with ChAdOx1 | 1,868,472 | 119,127 | 12 | 12 | 1.00 (0.57 to 1.76) |
| Venous thromboembolism narrow with thrombocytopenia 42 days pre to 14 days post | Vaccinated with BNT162b2 | 1,660,903 | 118,637 | 10 | 13 | 0.77 (0.42 to 1.44) |
| Venous thromboembolism narrow with thrombocytopenia 42 days pre to 14 days post | SARS-CoV-2 PCR positive test | 299,199 | 40,595 | 6 | 1.6 | 3.72 (1.67 to 8.29) |

### Results for vaccinated stratified by calendar month

For each event of interest the number of persons contributing to the analysis from the target population, their person-years contributed, and the number of events observed for them are given. Their expected events are estimated using indirect standardisation to the general population, with expected events giving the number of events we would have expected to have seen if their outcome experience was the same as that of the general population. Standardised incidence ratios (SIRs) with 95% confidence intervals (CIs) were estimated. Events with less than 5 occurrences have been omitted for privacy reasons.

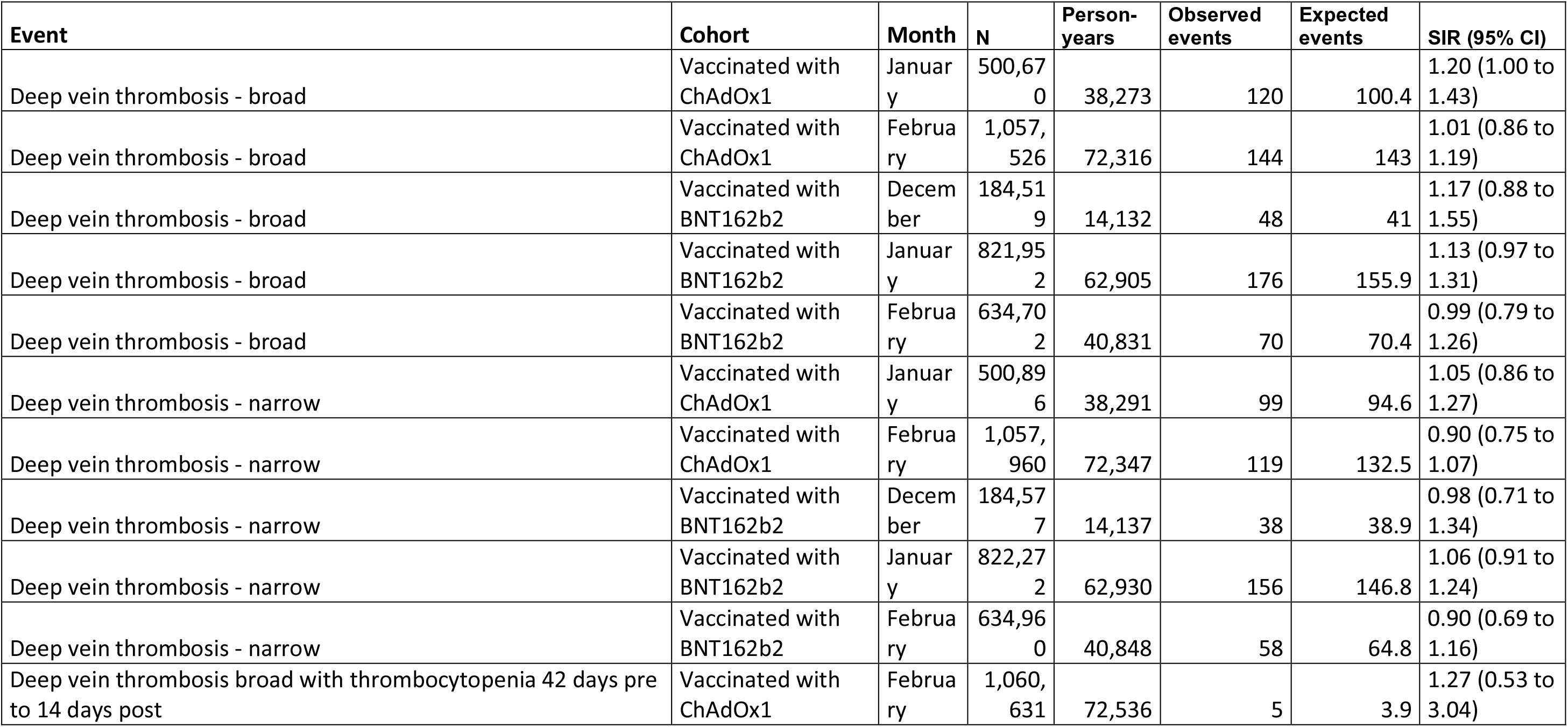
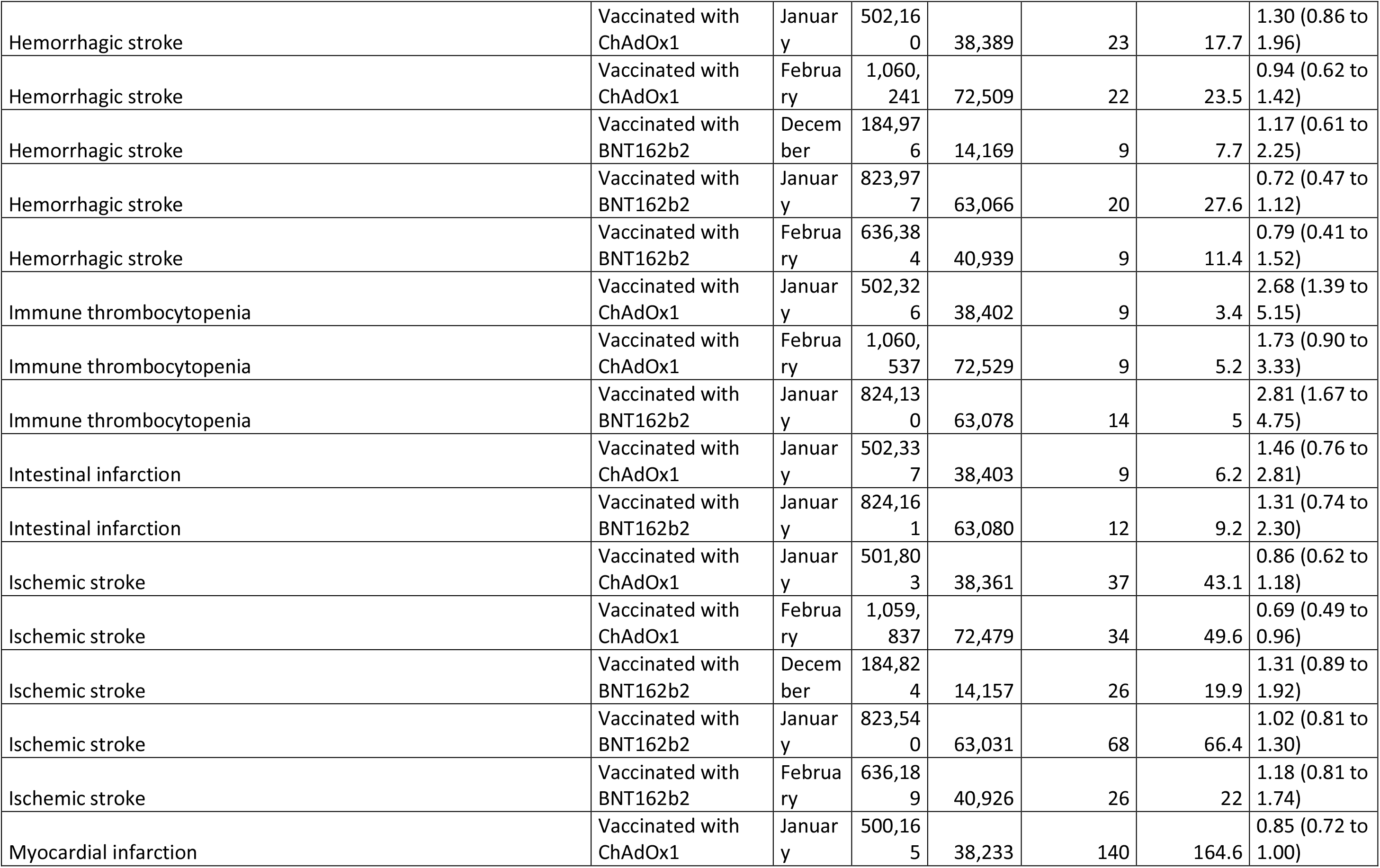
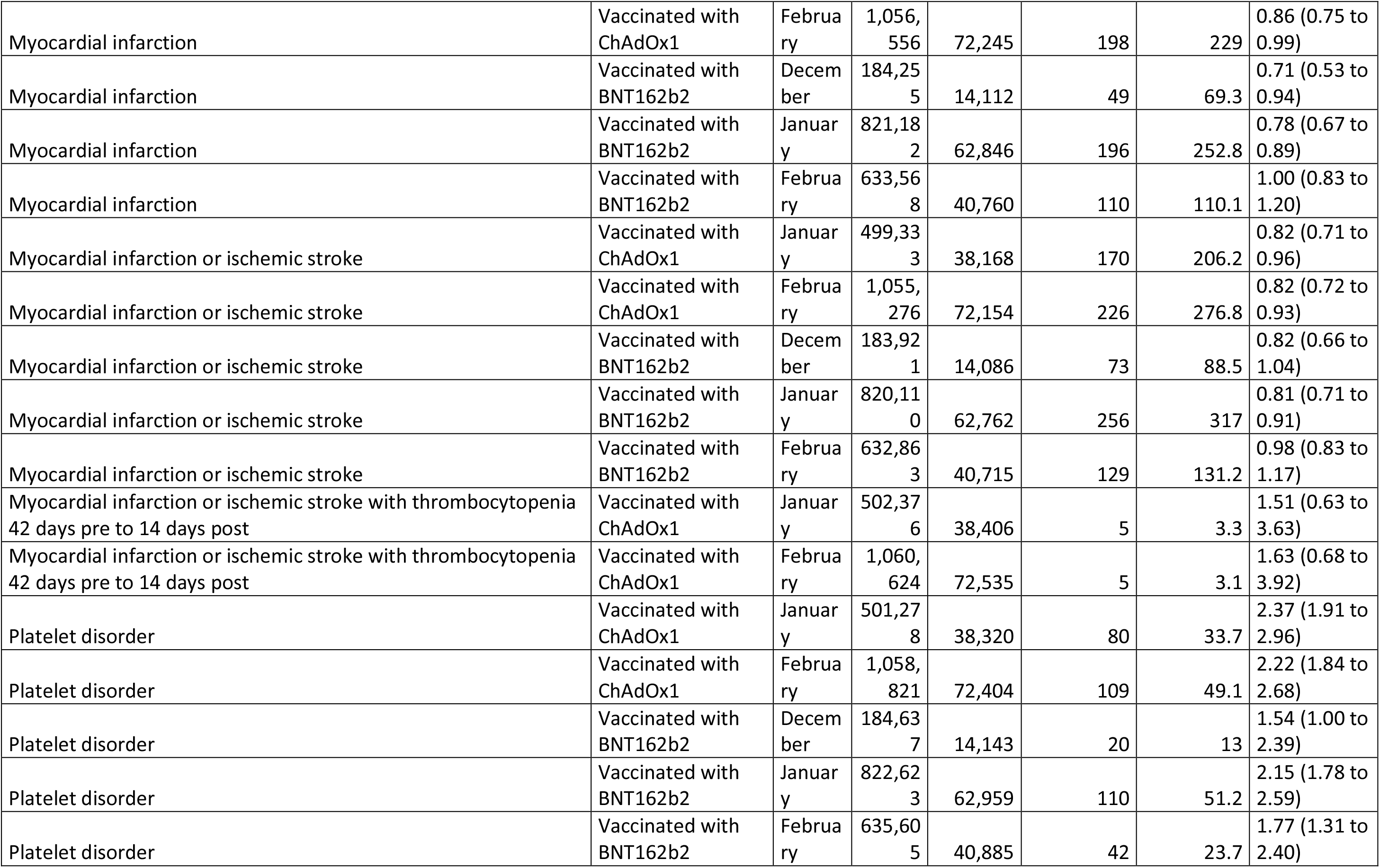
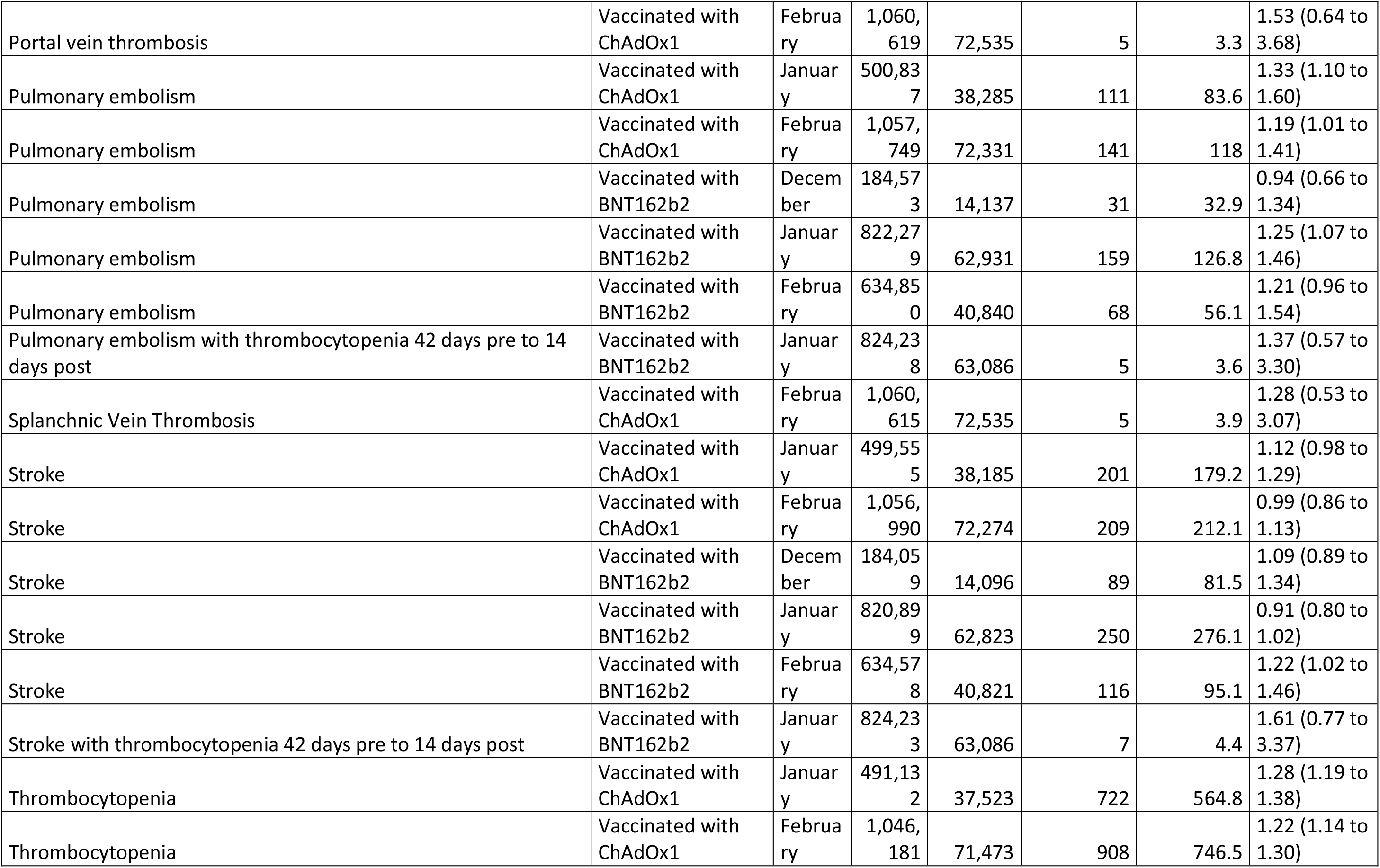
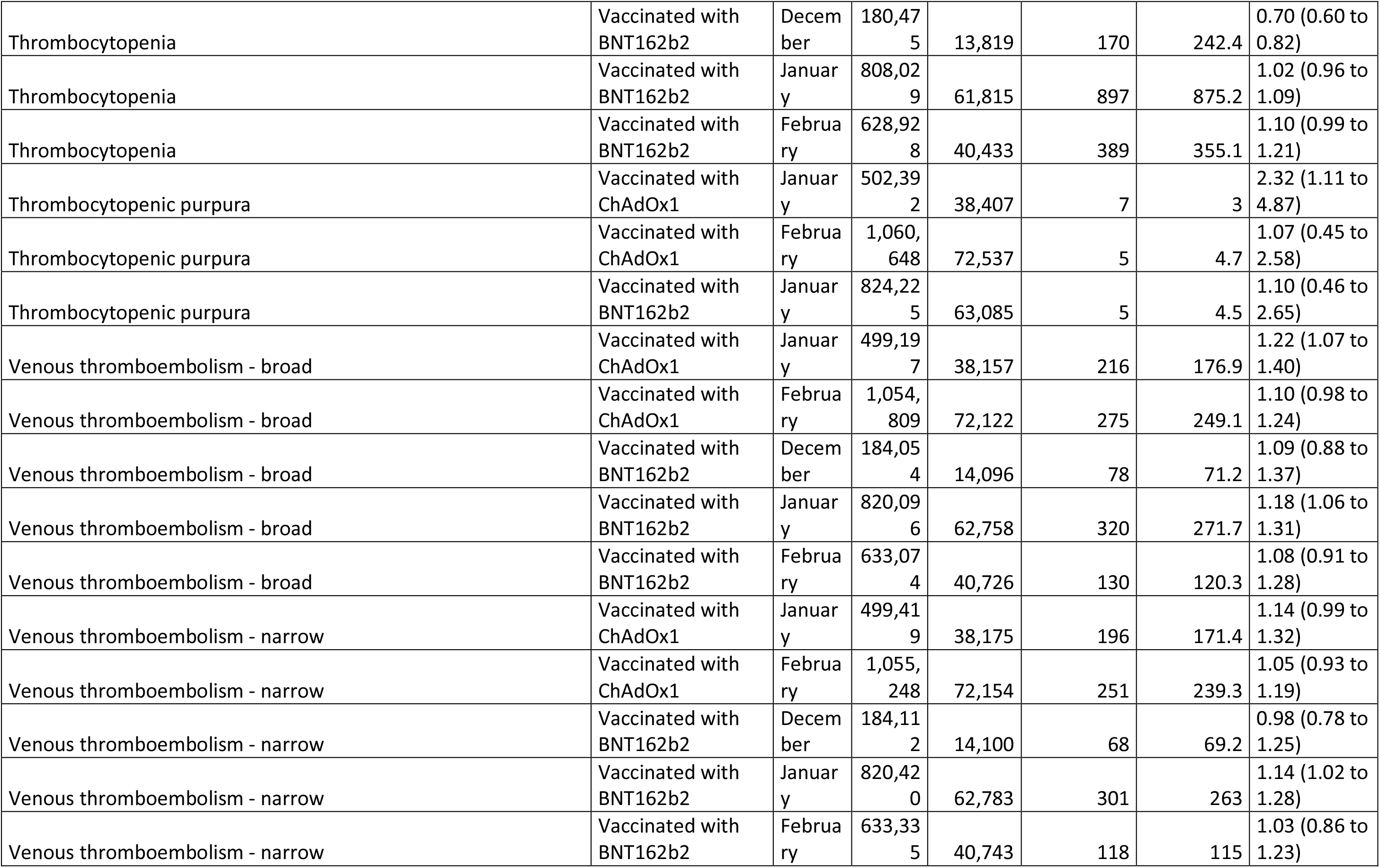
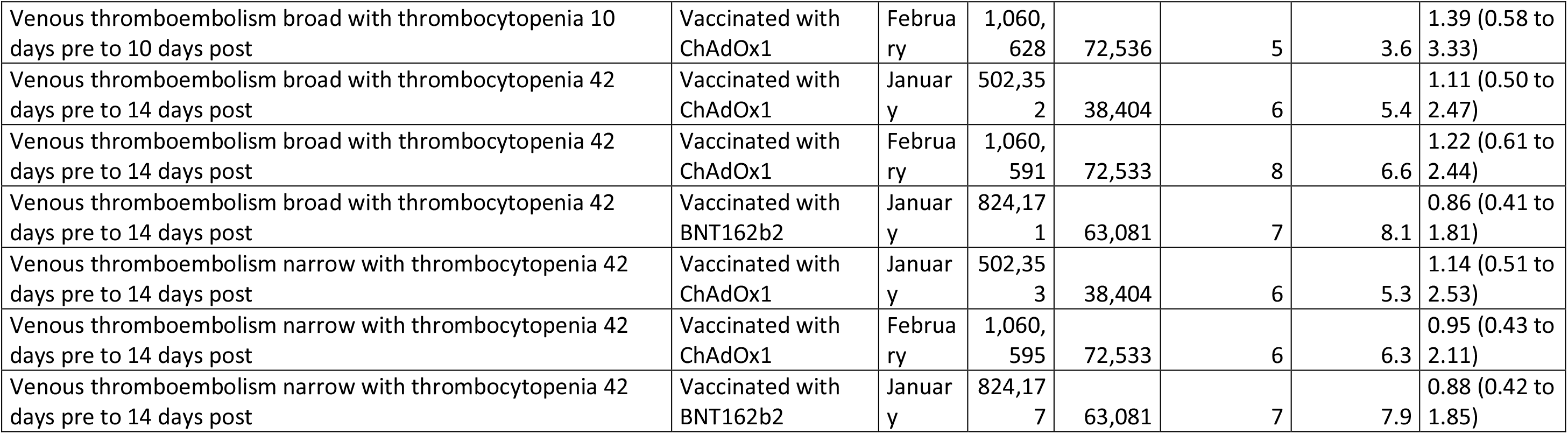

### Results without requiring year of prior history

For each event of interest the number of persons contributing to the analysis from the target population, their person-years contributed, and the number of events observed for them are given. Their expected events are estimated using indirect standardisation to the general population, with expected events giving the number of events we would have expected to have seen if their outcome experience was the same as that of the general population. Standardised incidence ratios (SIRs) with 95% confidence intervals (CIs) were estimated. Events with less than 5 occurrences have been omitted for privacy reasons.

| Event | Cohort | N | Person-years | Observed events | Expected events | SIR (95% CI) |
| --- | --- | --- | --- | --- | --- | --- |
| Cerebral venous sinus thrombosis | Vaccinated with ChAdOx1 | 1,956,136 | 124,569 | 5 | 2.2 | 2.32 (0.97 to 5.58) |
| Deep vein thrombosis - broad | Vaccinated with ChAdOx1 | 1,950,188 | 124,172 | 305 | 275.8 | 1.11 (0.99 to 1.24) |
| Deep vein thrombosis - broad | Vaccinated with BNT162b2 | 1,726,968 | 123,253 | 312 | 289.5 | 1.08 (0.96 to 1.20) |
| Deep vein thrombosis - broad | SARS-CoV-2 PCR positive test | 298,832 | 40,538 | 143 | 55.1 | 2.60 (2.20 to 3.06) |
| Deep vein thrombosis - narrow | Vaccinated with ChAdOx1 | 1,950,982 | 124,225 | 253 | 258 | 0.98 (0.87 to 1.11) |
| Deep vein thrombosis - narrow | Vaccinated with BNT162b2 | 1,727,622 | 123,301 | 268 | 272.3 | 0.98 (0.87 to 1.11) |
| Deep vein thrombosis - narrow | SARS-CoV-2 PCR positive test | 298,901 | 40,549 | 125 | 50.8 | 2.46 (2.07 to 2.93) |
| Deep vein thrombosis broad with thrombocytopenia 10 days pre to 10 days post | Vaccinated with ChAdOx1 | 1,956,124 | 124,567 | 9 | 4.6 | 1.96 (1.02 to 3.77) |
| Deep vein thrombosis broad with thrombocytopenia 42 days pre to 14 days post | Vaccinated with ChAdOx1 | 1,956,087 | 124,564 | 12 | 8 | 1.51 (0.86 to 2.65) |
| Deep vein thrombosis broad with thrombocytopenia 42 days pre to 14 days post | Vaccinated with BNT162b2 | 1,731,862 | 123,610 | 5 | 8.4 | 0.59 (0.25 to 1.42) |
| Deep vein thrombosis narrow with thrombocytopenia 10 days pre to 10 days post | Vaccinated with ChAdOx1 | 1,956,131 | 124,568 | 6 | 4.3 | 1.39 (0.62 to 3.09) |
| Deep vein thrombosis narrow with thrombocytopenia 42 days pre to 14 days post | Vaccinated with ChAdOx1 | 1,956,096 | 124,565 | 9 | 7.6 | 1.19 (0.62 to 2.29) |
| Deep vein thrombosis narrow with thrombocytopenia 42 days pre to 14 days post | Vaccinated with BNT162b2 | 1,731,870 | 123,610 | 5 | 8.1 | 0.62 (0.26 to 1.48) |
| Hemorrhagic stroke | Vaccinated with ChAdOx1 | 1,955,323 | 124,513 | 51 | 47.2 | 1.08 (0.82 to 1.42) |
| Hemorrhagic stroke | Vaccinated with BNT162b2 | 1,731,304 | 123,569 | 38 | 51.3 | 0.74 (0.54 to 1.02) |
| Hemorrhagic stroke | SARS-CoV-2 PCR positive test | 299,252 | 40,605 | 11 | 8.5 | 1.29 (0.71 to 2.33) |
| Immune thrombocytopenia | Vaccinated with ChAdOx1 | 1,955,913 | 124,552 | 18 | 9.9 | 1.82 (1.14 to 2.88) |
| Immune thrombocytopenia | Vaccinated with BNT162b2 | 1,731,680 | 123,596 | 16 | 9.5 | 1.68 (1.03 to 2.75) |
| Immune thrombocytopenia | SARS-CoV-2 PCR positive test | 299,300 | 40,614 | 5 | 1.8 | 2.71 (1.13 to 6.52) |
| Intestinal infarction | Vaccinated with ChAdOx1 | 1,955,897 | 124,551 | 16 | 15.8 | 1.01 (0.62 to 1.66) |
| Intestinal infarction | Vaccinated with BNT162b2 | 1,731,725 | 123,600 | 18 | 16 | 1.12 (0.71 to 1.78) |
| Ischemic stroke | Vaccinated with ChAdOx1 | 1,954,444 | 124,451 | 83 | 103.1 | 0.81 (0.65 to 1.00) |
| Ischemic stroke | Vaccinated with BNT162b2 | 1,730,486 | 123,507 | 124 | 115.7 | 1.07 (0.90 to 1.28) |
| Ischemic stroke | SARS-CoV-2 PCR positive test | 299,225 | 40,603 | 13 | 12.6 | 1.03 (0.60 to 1.77) |
| Myocardial infarction | Vaccinated with ChAdOx1 | 1,948,615 | 124,056 | 368 | 440.2 | 0.84 (0.75 to 0.93) |
| Myocardial infarction | Vaccinated with BNT162b2 | 1,724,692 | 123,097 | 375 | 459 | 0.82 (0.74 to 0.90) |
| Myocardial infarction | SARS-CoV-2 PCR positive test | 298,757 | 40,530 | 79 | 67.2 | 1.17 (0.94 to 1.46) |
| Myocardial infarction or ischemic stroke | Vaccinated with ChAdOx1 | 1,946,125 | 123,887 | 438 | 539.6 | 0.81 (0.74 to 0.89) |
| Myocardial infarction or ischemic stroke | Vaccinated with BNT162b2 | 1,722,509 | 122,937 | 481 | 570.7 | 0.84 (0.77 to 0.92) |
| Myocardial infarction or ischemic stroke | SARS-CoV-2 PCR positive test | 298,589 | 40,506 | 88 | 79.4 | 1.11 (0.90 to 1.37) |
| Myocardial infarction or ischemic stroke with thrombocytopenia 10 days pre to 10 days post | Vaccinated with ChAdOx1 | 1,956,120 | 124,567 | 5 | 3.6 | 1.39 (0.58 to 3.34) |
| Myocardial infarction or ischemic stroke with thrombocytopenia 10 days pre to 10 days post | Vaccinated with BNT162b2 | 1,731,873 | 123,611 | 5 | 4.5 | 1.12 (0.47 to 2.69) |
| Myocardial infarction or ischemic stroke with thrombocytopenia 42 days pre to 14 days post | Vaccinated with ChAdOx1 | 1,956,063 | 124,563 | 10 | 7.2 | 1.39 (0.75 to 2.58) |
| Myocardial infarction or ischemic stroke with thrombocytopenia 42 days pre to 14 days post | Vaccinated with BNT162b2 | 1,731,829 | 123,607 | 7 | 8.8 | 0.80 (0.38 to 1.67) |
| Platelet disorder | Vaccinated with ChAdOx1 | 1,952,742 | 124,328 | 216 | 94.4 | 2.29 (2.00 to 2.62) |
| Platelet disorder | Vaccinated with BNT162b2 | 1,728,759 | 123,378 | 184 | 95.6 | 1.93 (1.67 to 2.22) |
| Platelet disorder | SARS-CoV-2 PCR positive test | 299,047 | 40,574 | 57 | 16.8 | 3.40 (2.62 to 4.41) |
| Portal vein thrombosis | Vaccinated with ChAdOx1 | 1,956,077 | 124,565 | 8 | 5.7 | 1.41 (0.71 to 2.82) |
| Pulmonary embolism | Vaccinated with ChAdOx1 | 1,950,701 | 124,202 | 281 | 225.9 | 1.24 (1.11 to 1.40) |
| Pulmonary embolism | Vaccinated with BNT162b2 | 1,727,494 | 123,293 | 282 | 231.3 | 1.22 (1.09 to 1.37) |
| Pulmonary embolism | SARS-CoV-2 PCR positive test | 298,910 | 40,513 | 547 | 37 | 14.77 (13.58 to 16.06) |
| Pulmonary embolism with thrombocytopenia 42 days pre to 14 days post | Vaccinated with ChAdOx1 | 1,956,118 | 124,567 | 6 | 5.9 | 1.02 (0.46 to 2.26) |
| Pulmonary embolism with thrombocytopenia 42 days pre to 14 days post | Vaccinated with BNT162b2 | 1,731,900 | 123,612 | 7 | 6.1 | 1.15 (0.55 to 2.40) |
| Pulmonary embolism with thrombocytopenia 42 days pre to 14 days post | SARS-CoV-2 PCR positive test | 299,333 | 40,618 | 6 | 0.8 | 7.96 (3.58 to 17.71) |
| Splanchnic Vein Thrombosis | Vaccinated with ChAdOx1 | 1,956,072 | 124,564 | 9 | 6.9 | 1.31 (0.68 to 2.52) |
| Stroke | Vaccinated with ChAdOx1 | 1,948,412 | 124,026 | 481 | 443 | 1.09 (0.99 to 1.19) |
| Stroke | Vaccinated with BNT162b2 | 1,725,125 | 123,110 | 482 | 495.6 | 0.97 (0.89 to 1.06) |
| Stroke | SARS-CoV-2 PCR positive test | 298,834 | 40,547 | 66 | 56.9 | 1.16 (0.91 to 1.48) |
| Stroke with thrombocytopenia 10 days pre to 10 days post | Vaccinated with ChAdOx1 | 1,956,146 | 124,568 | 6 | 3.2 | 1.88 (0.85 to 4.19) |
| Stroke with thrombocytopenia 42 days pre to 14 days post | Vaccinated with ChAdOx1 | 1,956,114 | 124,566 | 7 | 6.5 | 1.07 (0.51 to 2.25) |
| Stroke with thrombocytopenia 42 days pre to 14 days post | Vaccinated with BNT162b2 | 1,731,882 | 123,611 | 8 | 7.7 | 1.04 (0.52 to 2.09) |
| Thrombocytopenia | Vaccinated with ChAdOx1 | 1,927,116 | 122,484 | 1,865 | 1,517.30 | 1.23 (1.17 to 1.29) |
| Thrombocytopenia | Vaccinated with BNT162b2 | 1,702,494 | 121,405 | 1,548 | 1,624.40 | 0.95 (0.91 to 1.00) |
| Thrombocytopenia | SARS-CoV-2 PCR positive test | 297,122 | 40,310 | 254 | 266.7 | 0.95 (0.84 to 1.08) |
| Thrombocytopenic purpura | Vaccinated with ChAdOx1 | 1,956,098 | 124,565 | 12 | 9.2 | 1.31 (0.74 to 2.31) |
| Thrombocytopenic purpura | Vaccinated with BNT162b2 | 1,731,847 | 123,609 | 6 | 8.9 | 0.68 (0.30 to 1.51) |
| Venous thromboembolism - broad | Vaccinated with ChAdOx1 | 1,945,125 | 123,830 | 558 | 479.9 | 1.16 (1.07 to 1.26) |
| Venous thromboembolism - broad | Vaccinated with BNT162b2 | 1,722,829 | 122,953 | 569 | 499.3 | 1.14 (1.05 to 1.24) |
| Venous thromboembolism - broad | SARS-CoV-2 PCR positive test | 298,453 | 40,439 | 664 | 87.3 | 7.61 (7.05 to 8.21) |
| Venous thromboembolism - narrow | Vaccinated with ChAdOx1 | 1,945,919 | 123,883 | 508 | 463.2 | 1.10 (1.01 to 1.20) |
| Venous thromboembolism - narrow | Vaccinated with BNT162b2 | 1,723,490 | 123,001 | 526 | 482.9 | 1.09 (1.00 to 1.19) |
| Venous thromboembolism - narrow | SARS-CoV-2 PCR positive test | 298,521 | 40,450 | 646 | 83.2 | 7.76 (7.19 to 8.39) |
| Venous thromboembolism broad with thrombocytopenia 10 days pre to 10 days post | Vaccinated with ChAdOx1 | 1,956,082 | 124,564 | 12 | 7.2 | 1.66 (0.94 to 2.92) |
| Venous thromboembolism broad with thrombocytopenia 10 days pre to 10 days post | Vaccinated with BNT162b2 | 1,731,858 | 123,609 | 7 | 7.6 | 0.92 (0.44 to 1.93) |
| Venous thromboembolism broad with thrombocytopenia 42 days pre to 14 days post | Vaccinated with ChAdOx1 | 1,956,004 | 124,558 | 17 | 13.2 | 1.29 (0.80 to 2.07) |
| Venous thromboembolism broad with thrombocytopenia 42 days pre to 14 days post | Vaccinated with BNT162b2 | 1,731,794 | 123,604 | 11 | 13.9 | 0.79 (0.44 to 1.42) |
| Venous thromboembolism broad with thrombocytopenia 42 days pre to 14 days post | SARS-CoV-2 PCR positive test | 299,324 | 40,617 | 6 | 1.7 | 3.46 (1.55 to 7.69) |
| Venous thromboembolism narrow with thrombocytopenia 10 days pre to 10 days post | Vaccinated with ChAdOx1 | 1,956,088 | 124,564 | 9 | 7 | 1.28 (0.67 to 2.47) |
| Venous thromboembolism narrow with thrombocytopenia 10 days pre to 10 days post | Vaccinated with BNT162b2 | 1,731,863 | 123,610 | 7 | 7.4 | 0.94 (0.45 to 1.98) |
| Venous thromboembolism narrow with thrombocytopenia 42 days pre to 14 days post | Vaccinated with ChAdOx1 | 1,956,011 | 124,559 | 14 | 12.9 | 1.09 (0.64 to 1.83) |
| Venous thromboembolism narrow with thrombocytopenia 42 days pre to 14 days post | Vaccinated with BNT162b2 | 1,731,801 | 123,605 | 11 | 13.6 | 0.81 (0.45 to 1.46) |
| Venous thromboembolism narrow with thrombocytopenia 42 days pre to 14 days post | SARS-CoV-2 PCR positive test | 299,324 | 40,617 | 6 | 1.7 | 3.59 (1.61 to 8.00) |

### Results with general population identified as of 1^st^ December 2017

For each event of interest the number of persons contributing to the analysis from the target population, their person-years contributed, and the number of events observed for them are given. Their expected events are estimated using indirect standardisation to the general population, with expected events giving the number of events we would have expected to have seen if their outcome experience was the same as that of the general population. Standardised incidence ratios (SIRs) with 95% confidence intervals (CIs) were estimated. Events with less than 5 occurrences have been omitted for privacy reasons. *Cerebral venous sinus thrombosis is reported without the requirement for a year of prior history as less than 5 events were seen when this restriction was imposed.

| Event | Cohort | N | Person-years | Observed events | Expected events | SIR (95% CI) |
| --- | --- | --- | --- | --- | --- | --- |
| Cerebral venous sinus thrombosis* | Vaccinated with ChAdOx1 | 1,956,136 | 124,569 | 5 | 1.8 | 2.70 (1.13 to 6.49) |
| Deep vein thrombosis - broad | Vaccinated with ChAdOx1 | 1,862,896 | 118,757 | 275 | 239.4 | 1.15 (1.02 to 1.29) |
| Deep vein thrombosis - broad | Vaccinated with BNT162b2 | 1,656,235 | 118,297 | 296 | 249.7 | 1.19 (1.06 to 1.33) |
| Deep vein thrombosis - broad | SARS-CoV-2 PCR positive test | 298,707 | 40,517 | 143 | 46.9 | 3.05 (2.59 to 3.59) |
| Deep vein thrombosis - narrow | Vaccinated with ChAdOx1 | 1,863,668 | 118,809 | 226 | 223.2 | 1.01 (0.89 to 1.15) |
| Deep vein thrombosis - narrow | Vaccinated with BNT162b2 | 1,656,875 | 118,344 | 254 | 234 | 1.09 (0.96 to 1.23) |
| Deep vein thrombosis - narrow | SARS-CoV-2 PCR positive test | 298,776 | 40,528 | 125 | 43 | 2.90 (2.44 to 3.46) |
| Deep vein thrombosis broad with thrombocytopenia 10 days pre to 10 days post | Vaccinated with ChAdOx1 | 1,868,584 | 119,135 | 6 | 3.9 | 1.55 (0.70 to 3.45) |
| Deep vein thrombosis broad with thrombocytopenia 42 days pre to 14 days post | Vaccinated with ChAdOx1 | 1,868,547 | 119,132 | 9 | 6.9 | 1.30 (0.68 to 2.51) |
| Deep vein thrombosis narrow with thrombocytopenia 42 days pre to 14 days post | Vaccinated with ChAdOx1 | 1,868,554 | 119,133 | 7 | 6.5 | 1.07 (0.51 to 2.25) |
| Hemorrhagic stroke | Vaccinated with ChAdOx1 | 1,867,822 | 119,084 | 45 | 40.9 | 1.10 (0.82 to 1.47) |
| Hemorrhagic stroke | Vaccinated with BNT162b2 | 1,660,431 | 118,603 | 38 | 44.3 | 0.86 (0.62 to 1.18) |
| Hemorrhagic stroke | SARS-CoV-2 PCR positive test | 299,127 | 40,584 | 11 | 7.5 | 1.47 (0.82 to 2.66) |
| Immune thrombocytopenia | Vaccinated with ChAdOx1 | 1,868,376 | 119,120 | 18 | 8.4 | 2.15 (1.35 to 3.41) |
| Immune thrombocytopenia | Vaccinated with BNT162b2 | 1,660,786 | 118,628 | 15 | 8 | 1.87 (1.13 to 3.11) |
| Immune thrombocytopenia | SARS-CoV-2 PCR positive test | 299,175 | 40,593 | 5 | 1.5 | 3.23 (1.35 to 7.77) |
| Intestinal infarction | Vaccinated with ChAdOx1 | 1,868,365 | 119,120 | 13 | 14.3 | 0.91 (0.53 to 1.56) |
| Intestinal infarction | Vaccinated with BNT162b2 | 1,660,825 | 118,631 | 17 | 14.6 | 1.16 (0.72 to 1.87) |
| Ischemic stroke | Vaccinated with ChAdOx1 | 1,867,000 | 119,025 | 72 | 91.2 | 0.79 (0.63 to 0.99) |
| Ischemic stroke | Vaccinated with BNT162b2 | 1,659,635 | 118,542 | 120 | 102.6 | 1.17 (0.98 to 1.40) |
| Ischemic stroke | SARS-CoV-2 PCR positive test | 299,100 | 40,582 | 13 | 11.4 | 1.14 (0.66 to 1.96) |
| Myocardial infarction | Vaccinated with ChAdOx1 | 1,861,365 | 118,643 | 347 | 391.6 | 0.89 (0.80 to 0.98) |
| Myocardial infarction | Vaccinated with BNT162b2 | 1,654,037 | 118,145 | 356 | 409.8 | 0.87 (0.78 to 0.96) |
| Myocardial infarction | SARS-CoV-2 PCR positive test | 298,632 | 40,509 | 79 | 61.1 | 1.29 (1.04 to 1.61) |
| Myocardial infarction or ischemic stroke | Vaccinated with ChAdOx1 | 1,858,974 | 118,481 | 406 | 479.5 | 0.85 (0.77 to 0.93) |
| Myocardial infarction or ischemic stroke | Vaccinated with BNT162b2 | 1,651,903 | 117,990 | 458 | 508.9 | 0.90 (0.82 to 0.99) |
| Myocardial infarction or ischemic stroke | SARS-CoV-2 PCR positive test | 298,464 | 40,485 | 88 | 72.1 | 1.22 (0.99 to 1.50) |
| Myocardial infarction or ischemic stroke with thrombocytopenia 10 days pre to 10 days post | Vaccinated with ChAdOx1 | 1,868,580 | 119,135 | 5 | 3 | 1.66 (0.69 to 3.99) |
| Myocardial infarction or ischemic stroke with thrombocytopenia 42 days pre to 14 days post | Vaccinated with ChAdOx1 | 1,868,524 | 119,131 | 10 | 6.2 | 1.62 (0.87 to 3.02) |
| Myocardial infarction or ischemic stroke with thrombocytopenia 42 days pre to 14 days post | Vaccinated with BNT162b2 | 1,660,931 | 118,639 | 6 | 7.8 | 0.77 (0.35 to 1.71) |
| Platelet disorder | Vaccinated with ChAdOx1 | 1,865,323 | 118,905 | 198 | 81.2 | 2.44 (2.12 to 2.80) |
| Platelet disorder | Vaccinated with BNT162b2 | 1,657,943 | 118,416 | 174 | 82 | 2.12 (1.83 to 2.46) |
| Platelet disorder | SARS-CoV-2 PCR positive test | 298,922 | 40,552 | 57 | 14 | 4.08 (3.14 to 5.28) |
| Portal vein thrombosis | Vaccinated with ChAdOx1 | 1,868,539 | 119,133 | 8 | 5.2 | 1.55 (0.77 to 3.09) |
| Pulmonary embolism | Vaccinated with ChAdOx1 | 1,863,403 | 118,787 | 259 | 198.7 | 1.30 (1.15 to 1.47) |
| Pulmonary embolism | Vaccinated with BNT162b2 | 1,656,755 | 118,336 | 261 | 202.8 | 1.29 (1.14 to 1.45) |
| Pulmonary embolism | SARS-CoV-2 PCR positive test | 298,786 | 40,491 | 547 | 32 | 17.09 (15.72 to 18.58) |
| Pulmonary embolism with thrombocytopenia 42 days pre to 14 days post | Vaccinated with ChAdOx1 | 1,868,576 | 119,134 | 6 | 5.2 | 1.16 (0.52 to 2.58) |
| Pulmonary embolism with thrombocytopenia 42 days pre to 14 days post | Vaccinated with BNT162b2 | 1,660,999 | 118,644 | 7 | 5.5 | 1.27 (0.60 to 2.65) |
| Pulmonary embolism with thrombocytopenia 42 days pre to 14 days post | SARS-CoV-2 PCR positive test | 299,208 | 40,596 | 6 | 0.7 | 9.05 (4.07 to 20.14) |
| Splanchnic Vein Thrombosis | Vaccinated with ChAdOx1 | 1,868,536 | 119,132 | 9 | 6.2 | 1.46 (0.76 to 2.80) |
| Stroke | Vaccinated with ChAdOx1 | 1,861,358 | 118,629 | 413 | 385.8 | 1.07 (0.97 to 1.18) |
| Stroke | Vaccinated with BNT162b2 | 1,654,569 | 118,167 | 457 | 429.2 | 1.06 (0.97 to 1.17) |
| Stroke | SARS-CoV-2 PCR positive test | 298,709 | 40,525 | 65 | 50.4 | 1.29 (1.01 to 1.64) |
| Stroke with thrombocytopenia 10 days pre to 10 days post | Vaccinated with ChAdOx1 | 1,868,605 | 119,136 | 6 | 2.6 | 2.32 (1.04 to 5.16) |
| Stroke with thrombocytopenia 42 days pre to 14 days post | Vaccinated with ChAdOx1 | 1,868,573 | 119,134 | 6 | 5.5 | 1.09 (0.49 to 2.43) |
| Stroke with thrombocytopenia 42 days pre to 14 days post | Vaccinated with BNT162b2 | 1,660,982 | 118,643 | 8 | 6.5 | 1.24 (0.62 to 2.47) |
| Thrombocytopenia | Vaccinated with ChAdOx1 | 1,840,495 | 117,121 | 1,707 | 1,281.00 | 1.33 (1.27 to 1.40) |
| Thrombocytopenia | Vaccinated with BNT162b2 | 1,632,328 | 116,491 | 1,462 | 1,372.60 | 1.07 (1.01 to 1.12) |
| Thrombocytopenia | SARS-CoV-2 PCR positive test | 296,999 | 40,289 | 254 | 218.9 | 1.16 (1.03 to 1.31) |
| Thrombocytopenic purpura | Vaccinated with ChAdOx1 | 1,868,562 | 119,134 | 12 | 7.5 | 1.59 (0.90 to 2.80) |
| Thrombocytopenic purpura | Vaccinated with BNT162b2 | 1,660,955 | 118,641 | 5 | 7.3 | 0.68 (0.28 to 1.64) |
| Venous thromboembolism - broad | Vaccinated with ChAdOx1 | 1,858,064 | 118,431 | 508 | 419.1 | 1.21 (1.11 to 1.32) |
| Venous thromboembolism - broad | Vaccinated with BNT162b2 | 1,652,244 | 118,007 | 532 | 433.9 | 1.23 (1.13 to 1.33) |
| Venous thromboembolism - broad | SARS-CoV-2 PCR positive test | 298,329 | 40,418 | 664 | 74.8 | 8.88 (8.23 to 9.58) |
| Venous thromboembolism - narrow | Vaccinated with ChAdOx1 | 1,858,837 | 118,483 | 461 | 403.9 | 1.14 (1.04 to 1.25) |
| Venous thromboembolism - narrow | Vaccinated with BNT162b2 | 1,652,892 | 118,054 | 491 | 419 | 1.17 (1.07 to 1.28) |
| Venous thromboembolism - narrow | SARS-CoV-2 PCR positive test | 298,397 | 40,429 | 646 | 71.1 | 9.09 (8.42 to 9.82) |
| Venous thromboembolism broad with thrombocytopenia 10 days pre to 10 days post | Vaccinated with ChAdOx1 | 1,868,543 | 119,132 | 9 | 6.2 | 1.44 (0.75 to 2.77) |
| Venous thromboembolism broad with thrombocytopenia 10 days pre to 10 days post | Vaccinated with BNT162b2 | 1,660,959 | 118,641 | 6 | 6.7 | 0.90 (0.41 to 2.01) |
| Venous thromboembolism broad with thrombocytopenia 42 days pre to 14 days post | Vaccinated with ChAdOx1 | 1,868,467 | 119,126 | 14 | 11.6 | 1.21 (0.72 to 2.04) |
| Venous thromboembolism broad with thrombocytopenia 42 days pre to 14 days post | Vaccinated with BNT162b2 | 1,660,896 | 118,636 | 10 | 12.4 | 0.81 (0.43 to 1.50) |
| Venous thromboembolism broad with thrombocytopenia 42 days pre to 14 days post | SARS-CoV-2 PCR positive test | 299,199 | 40,595 | 6 | 1.5 | 3.93 (1.77 to 8.76) |
| Venous thromboembolism narrow with thrombocytopenia 10 days pre to 10 days post | Vaccinated with ChAdOx1 | 1,868,547 | 119,132 | 7 | 6 | 1.16 (0.56 to 2.44) |
| Venous thromboembolism narrow with thrombocytopenia 10 days pre to 10 days post | Vaccinated with BNT162b2 | 1,660,964 | 118,641 | 6 | 6.5 | 0.93 (0.42 to 2.06) |
| Venous thromboembolism narrow with thrombocytopenia 42 days pre to 14 days post | Vaccinated with ChAdOx1 | 1,868,472 | 119,127 | 12 | 11.3 | 1.07 (0.61 to 1.88) |
| Venous thromboembolism narrow with thrombocytopenia 42 days pre to 14 days post | Vaccinated with BNT162b2 | 1,660,903 | 118,637 | 10 | 12.1 | 0.83 (0.44 to 1.54) |
| Venous thromboembolism narrow with thrombocytopenia 42 days pre to 14 days post | SARS-CoV-2 PCR positive test | 299,199 | 40,595 | 6 | 1.5 | 4.11 (1.85 to 9.14) |

### Incidence rate ratios (IRRs) and 95% confidence intervals for pulmonary embolism stratified by age, sex, and calendar month

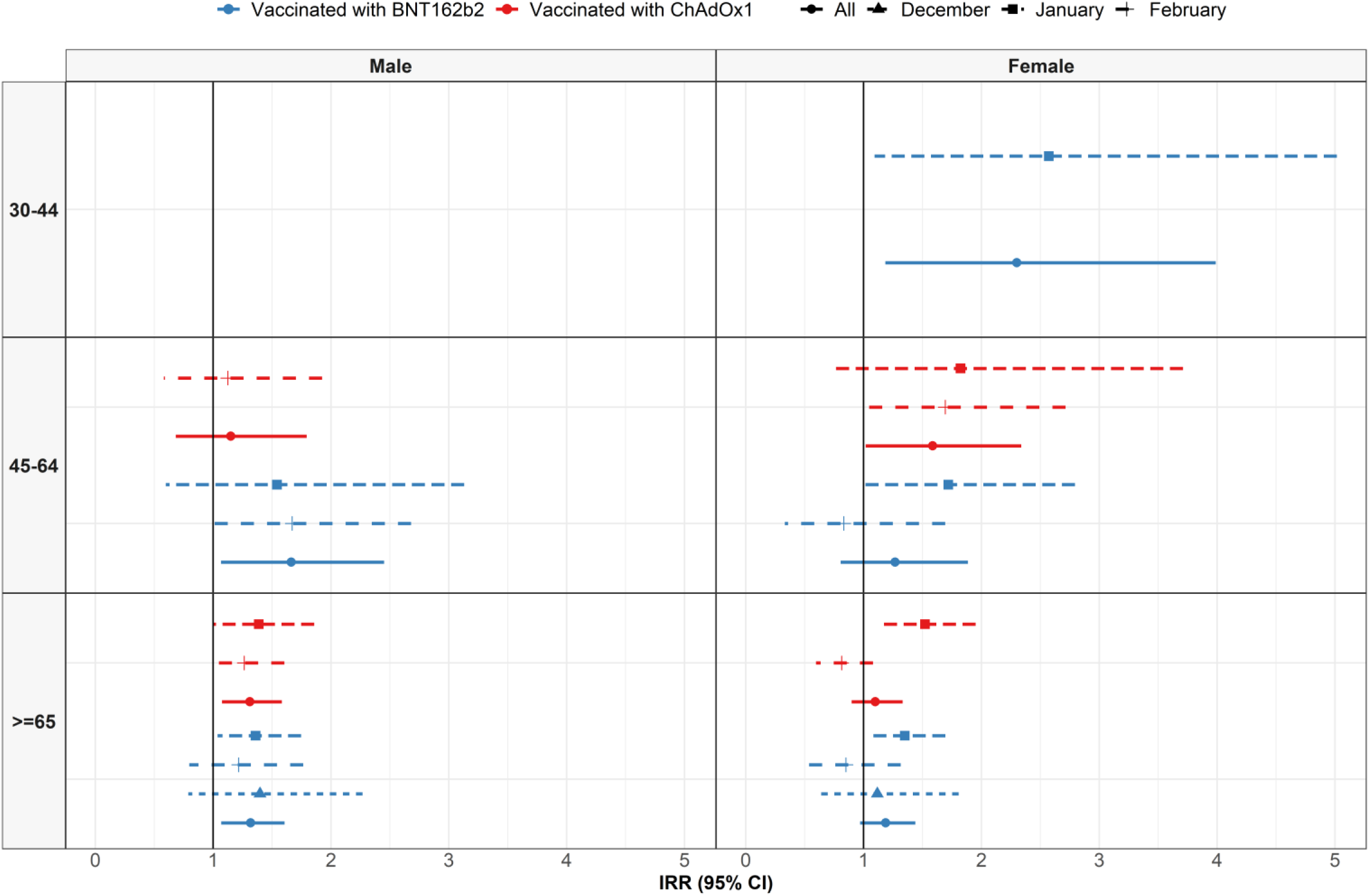

### Patient profiles: pulmonary embolism

The characteristics of persons with pulmonary embolism used for the primary analyses. *Conditions of interest: autoimmune disease, antiphospholipid syndrome, thrombophilia, asthma, atrial fibrillation, malignant neoplastic disease, diabetes mellitus, obesity, or renal impairment. ^†^Medications of interest included non-steroidal anti-inflammatory drugs, Cox2 inhibitors, systemic corticosteroids, hormonal contraceptives, tamoxifen, and sex hormones and modulators of the genital system

|  | General population | Vaccinated with ChAdOx1 | Vaccinated with BNT162b2 |
| --- | --- | --- | --- |
| N | 7,414 | 281 | 282 |
| Age | 68 [56 to 77] | 74 [67 to 79] | 75 [66 to 82] |
| Age: 30 to 39 | 474 (6.4%) | <5 | 6 (2.1%) |
| Age: 40 to 49 | 735 (9.9%) | 12 (4.3%) | 14 (5.0%) |
| Age: 50 to 59 | 1,164 (15.7%) | 30 (10.7%) | 23 (8.2%) |
| Age: 60 to 69 | 1,666 (22.5%) | 49 (17.4%) | 51 (18.1%) |
| Age: 70 to 79 | 1,918 (25.9%) | 119 (42.3%) | 89 (31.6%) |
| Age: 80 or older | 1,457 (19.7%) | 69 (24.6%) | 99 (35.1%) |
| Sex: Male | 3,543 (47.8%) | 138 (49.1%) | 135 (47.9%) |
| Years of prior observation time | 14.9 [7.9 to 17.6] | 17.8 [6.0 to 32.5] | 21.5 [8.7 to 34.9] |
| Comorbidities |  |  |  |
| Autoimmune disease | 325 (4.4%) | 13 (4.6%) | 18 (6.4%) |
| Antiphospholipid syndrome | 14 (0.2%) | 0 (0.0%) | 0 (0.0%) |
| Thrombophilia | 36 (0.5%) | 0 (0.0%) | 0 (0.0%) |
| Asthma | 1,137 (15.3%) | 43 (15.3%) | 39 (13.8%) |
| Atrial fibrillation | 302 (4.1%) | 19 (6.8%) | 14 (5.0%) |
| Malignant neoplastic disease | 1,533 (20.7%) | 91 (32.4%) | 94 (33.3%) |
| Diabetes mellitus | 974 (13.1%) | 51 (18.1%) | 46 (16.3%) |
| Obesity | 556 (7.5%) | 23 (8.2%) | 21 (7.4%) |
| Heart disease | 1,382 (18.6%) | 73 (26.0%) | 64 (22.7%) |
| Hypertensive disorder | 2,832 (38.2%) | 120 (42.7%) | 129 (45.7%) |
| Renal impairment | 1,131 (15.3%) | 63 (22.4%) | 71 (25.2%) |
| COPD | 674 (9.1%) | 31 (11.0%) | 33 (11.7%) |
| Dementia | 176 (2.4%) | 23 (8.2%) | 11 (3.9%) |
| Medication use (183 days prior to four days prior) |  |  |  |
| Non-steroidal anti-inflammatory drugs | 4,302 (58.0%) | 87 (31.0%) | 77 (27.3%) |
| Cox2 inhibitors | 42 (0.6%) | 0 (0.0%) | <5 |
| Systemic corticosteroids | 2,001 (27.0%) | 43 (15.3%) | 34 (12.1%) |
| Antithrombotic and anticoagulant therapies | 1,555 (21.0%) | 35 (12.5%) | 32 (11.3%) |
| Lipid modifying agents | 890 (12.0%) | 29 (10.3%) | 23 (8.2%) |
| Antineoplastic and immunomodulating agents | 387 (5.2%) | 11 (3.9%) | 12 (4.3%) |
| Hormonal contraceptives for systemic use | 158 (2.1%) | <5 | 6 (2.1%) |
| Tamoxifen | 39 (0.5%) | <5 | <5 |
| Sex hormones and modulators of the genital system | 295 (4.0%) | 9 (3.2%) | 9 (3.2%) |
| One or more condition of interest* | 3,388 (45.7%) | 165 (58.7%) | 176 (62.4%) |
| One or more medication of interest<sup>†</sup> | 4,746 (64.0%) | 108 (38.4%) | 100 (35.5%) |
| One or more condition/ medication of interest*<sup>†</sup> | 5,745 (77.5%) | 199 (70.8%) | 207 (73.4%) |

### Incidence rate ratios (IRRs) and 95% confidence intervals for thrombocytopenia stratified by age, sex, and calendar month

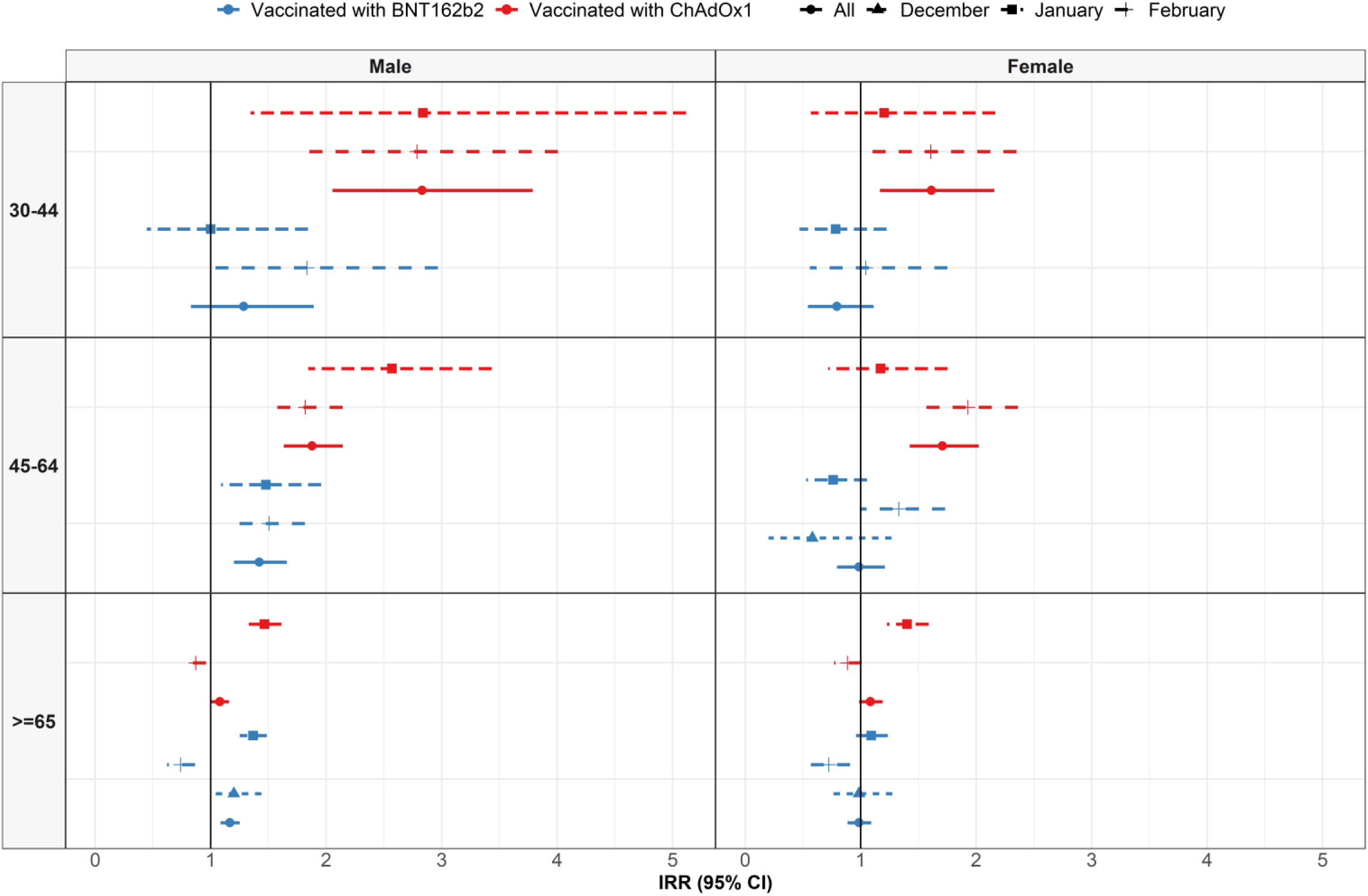

### Patient profiles: thrombocytopenia

The characteristics of persons with thrombocytopenia used for the primary analyses. *Conditions of interest: autoimmune disease, antiphospholipid syndrome, thrombophilia, asthma, atrial fibrillation, malignant neoplastic disease, diabetes mellitus, obesity, or renal impairment. ^†^Medications of interest included non-steroidal anti-inflammatory drugs, Cox2 inhibitors, systemic corticosteroids, hormonal contraceptives, tamoxifen, and sex hormones and modulators of the genital system

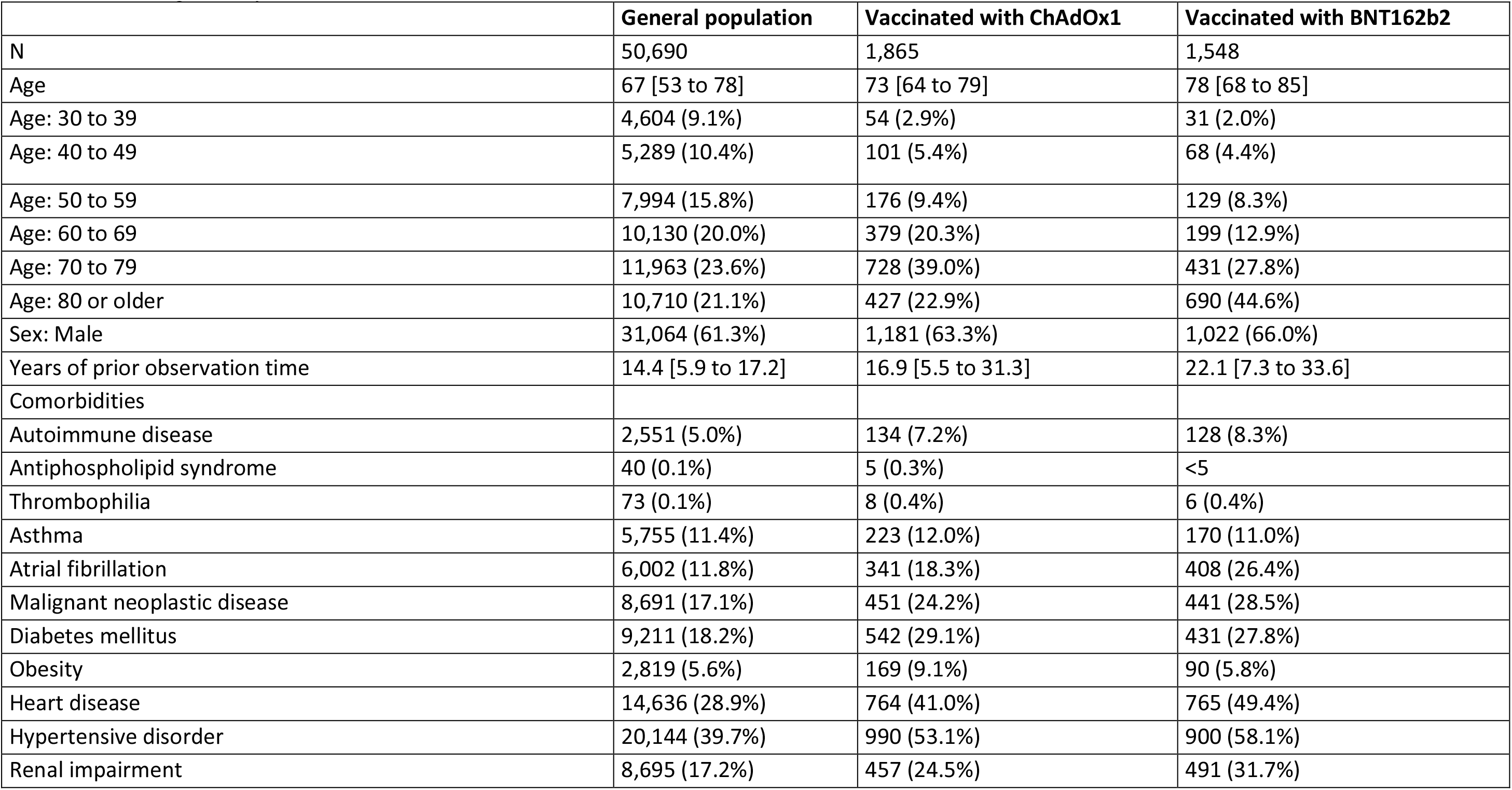
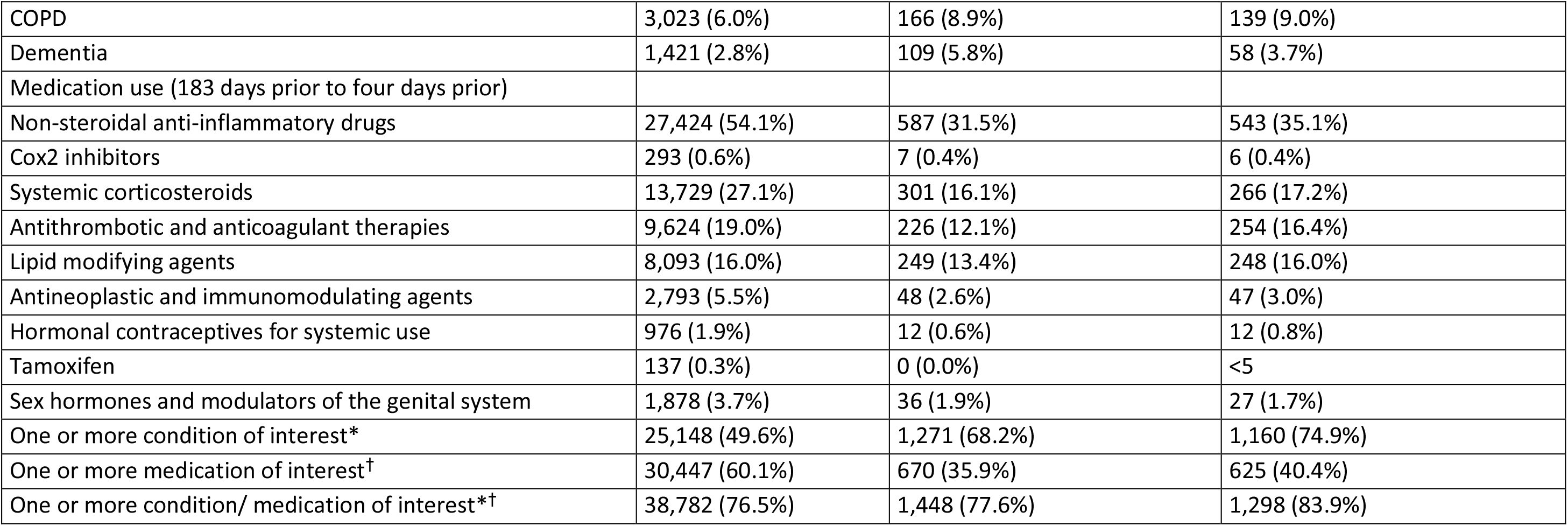

|  | General population | Vaccinated with ChAdOx1 | Vaccinated with BNT162b2 |
| --- | --- | --- | --- |
| N | 50,690 | 1,865 | 1,548 |
| Age | 67 [53 to 78] | 73 [64 to 79] | 78 [68 to 85] |
| Age: 30 to 39 | 4,604 (9.1%) | 54 (2.9%) | 31 (2.0%) |
| Age: 40 to 49 | 5,289 (10.4%) | 101 (5.4%) | 68 (4.4%) |
| Age: 50 to 59 | 7,994 (15.8%) | 176 (9.4%) | 129 (8.3%) |
| Age: 60 to 69 | 10,130 (20.0%) | 379 (20.3%) | 199 (12.9%) |
| Age: 70 to 79 | 11,963 (23.6%) | 728 (39.0%) | 431 (27.8%) |
| Age: 80 or older | 10,710 (21.1%) | 427 (22.9%) | 690 (44.6%) |
| Sex: Male | 31,064 (61.3%) | 1,181 (63.3%) | 1,022 (66.0%) |
| Years of prior observation time | 14.4 [5.9 to 17.2] | 16.9 [5.5 to 31.3] | 22.1 [7.3 to 33.6] |
| Comorbidities |  |  |  |
| Autoimmune disease | 2,551 (5.0%) | 134 (7.2%) | 128 (8.3%) |
| Antiphospholipid syndrome | 40 (0.1%) | 5 (0.3%) | <5 |
| Thrombophilia | 73 (0.1%) | 8 (0.4%) | 6 (0.4%) |
| Asthma | 5,755 (11.4%) | 223 (12.0%) | 170 (11.0%) |
| Atrial fibrillation | 6,002 (11.8%) | 341 (18.3%) | 408 (26.4%) |
| Malignant neoplastic disease | 8,691 (17.1%) | 451 (24.2%) | 441 (28.5%) |
| Diabetes mellitus | 9,211 (18.2%) | 542 (29.1%) | 431 (27.8%) |
| Obesity | 2,819 (5.6%) | 169 (9.1%) | 90 (5.8%) |
| Heart disease | 14,636 (28.9%) | 764 (41.0%) | 765 (49.4%) |
| Hypertensive disorder | 20,144 (39.7%) | 990 (53.1%) | 900 (58.1%) |
| Renal impairment | 8,695 (17.2%) | 457 (24.5%) | 491 (31.7%) |
| COPD | 3,023 (6.0%) | 166 (8.9%) | 139 (9.0%) |
| Dementia | 1,421 (2.8%) | 109 (5.8%) | 58 (3.7%) |
| Medication use (183 days prior to four days prior) |  |  |  |
| Non-steroidal anti-inflammatory drugs | 27,424 (54.1%) | 587 (31.5%) | 543 (35.1%) |
| Cox2 inhibitors | 293 (0.6%) | 7 (0.4%) | 6 (0.4%) |
| Systemic corticosteroids | 13,729 (27.1%) | 301 (16.1%) | 266 (17.2%) |
| Antithrombotic and anticoagulant therapies | 9,624 (19.0%) | 226 (12.1%) | 254 (16.4%) |
| Lipid modifying agents | 8,093 (16.0%) | 249 (13.4%) | 248 (16.0%) |
| Antineoplastic and immunomodulating agents | 2,793 (5.5%) | 48 (2.6%) | 47 (3.0%) |
| Hormonal contraceptives for systemic use | 976 (1.9%) | 12 (0.6%) | 12 (0.8%) |
| Tamoxifen | 137 (0.3%) | 0 (0.0%) | <5 |
| Sex hormones and modulators of the genital system | 1,878 (3.7%) | 36 (1.9%) | 27 (1.7%) |
| One or more condition of interest* | 25,148 (49.6%) | 1,271 (68.2%) | 1,160 (74.9%) |
| One or more medication of interest<sup>†</sup> | 30,447 (60.1%) | 670 (35.9%) | 625 (40.4%) |
| One or more condition/ medication of interest*<sup>†</sup> | 38,782 (76.5%) | 1,448 (77.6%) | 1,298 (83.9%) |

